# Outcome of COVID-19 in hospitalised immunocompromised patients: an analysis of the WHO ISARIC CCP-UK prospective cohort study

**DOI:** 10.1101/2022.08.08.22278576

**Authors:** Lance Turtle, Mathew Thorpe, Thomas M Drake, Maaike Swets, Carlo Palmieri, Clark D Russell, Antonia Ho, Stephen Aston, Daniel G Wootton, Alex Richter, Thushan I de Silva, Hayley E Hardwick, Gary Leeming, Andy Law, Peter JM Openshaw, Ewen M Harrison, ISARIC4C investigators, J Kenneth Baillie, Malcolm G Semple, Annemarie B Docherty

**Author notes:** **Corresponding author:** Lance Turtle, Ronald Ross Building, Department of Clinical Infection, Microbiology and Immunology, University of Liverpool, 8 West Derby Street, L69 7BE, Liverpool, UK. Equal contribution (Lance Turtle, Mathew Thorpe). Joint senior author (J Kenneth Baillie, Malcolm G Semple, Annemarie B Docherty).

## Abstract

**Background:** Immunocompromised patients may be at higher risk of mortality if hospitalised with COVID-19 compared with immunocompetent patients. However, previous studies have been contradictory. We aimed to determine whether immunocompromised patients were at greater risk of in-hospital death, and how this risk changed over the pandemic.

**Methods:** We included patients >=19yrs with symptomatic community-acquired COVID-19 recruited to the ISARIC WHO Clinical Characterisation Protocol UK. We defined immunocompromise as: immunosuppressant medication preadmission, cancer treatment, organ transplant, HIV, or congenital immunodeficiency. We used logistic regression to compare the risk of death in both groups, adjusting for age, sex, deprivation, ethnicity, vaccination and co-morbidities. We used Bayesian logistic regression to explore mortality over time.

**Findings:** Between 17/01/2020 and 28/02/2022 we recruited 156,552 eligible patients, of whom 21,954 (14%) were immunocompromised. 29% (n=6,499) of immunocompromised and 21% (n=28,608) of immunocompetent patients died in hospital. The odds of in-hospital mortality were elevated for immunocompromised patients (adjOR 1.44, 95% CI 1.39-1.50, p<0.001). As the pandemic progressed, in-hospital mortality reduced more slowly for immunocompromised patients than for immunocompetent patients. This was particularly evident with increasing age: the probability of the reduction in hospital mortality being less for immunocompromised patients aged 50-69yrs was 88% for men and 83% for women, and for those >80yrs was 99% for men, and 98% for women.

**Conclusions:** Immunocompromised patients remain at elevated risk of death from COVID-19. Targeted measures such as additional vaccine doses and monoclonal antibodies should be considered for this group.

**Funding:** National Institute for Health Research; Medical Research Council; Chief Scientist Office, Scotland.

## Introduction

Coronavirus disease 2019 (COVID-19), the disease caused by severe acute respiratory syndrome coronavirus 2 (SARS-CoV-2), disproportionately affects older people and those with underlying health conditions (1). However, a key challenge throughout the pandemic has been to delineate which comorbidities confer the greatest risk. Severe COVID-19 is an inflammatory process (2, 3), with several lines of evidence suggesting that immune dysfunction is linked to adverse outcome (4, 5). Consistent with this are clinical trial findings that anti-inflammatory treatments with dexamethasone, tocilizumab and baricitinib improve survival in patients with respiratory failure (6-8).

Early data in the pandemic suggested a high mortality among immunocompromised patients, with organ transplant recipients being at particular risk (9, 10). Studies have compared mortality among immunocompromised patients with other patient groups with COVID-19 with conflicting results. Some studies show increased mortality (11, 12), whereas others show no difference from other patient groups (13, 14). A challenge when attributing risk to individual comorbidities has been that many factors are correlated. For example, some kidney transplant recipients also have abnormal renal function which is itself a risk factor for poor outcome from COVID-19 (1, 15). A large UK population study using routine health data from 17 million primary care health records (OPENSafely) found that immunocompromising conditions, including organ transplant and haematological malignancy, increased the risk of COVID-19 associated death (16). However, detailed information on disease severity at presentation and events during hospitalisation were not available.

As public health measures to control the spread of SARS-CoV-2 have now eased, concern remains for the safety of those who are immunocompromised and whose vaccination response may be compromised (17). Our aim was therefore to analyse one of the largest prospective cohorts of hospitalised COVID-19 cases, the ISARIC WHO CCP-UK study dataset, to determine whether outcomes are worse in immunocompromised patients, and whether the improved survival observed in general over the course of the pandemic differed between immunocompromised and immunocompetent patients.

## Methods

### Study design and setting

The ISARIC WHO Clinical Characterisation Protocol in the United Kingdom (CCP-UK) was activated on 17th January 2020 as part of the public health response to the COVID-19 pandemic. CCP-UK prospectively recruited a cohort of >300,000 patients, hospitalised with COVID-19, from 306 healthcare facilities across the UK. The sample size was not pre-specified. The protocol, revision history, case report forms and consent forms, are available online at isaric4c.net. The study received ethical approval from the South Central - Oxford C Research Ethics Committee in England (Ref: 13/SC/0149) and by the Scotland A Research Ethics Committee (Ref: 20/SS/0028). This study is reported according to the Strengthening the Reporting of Observational Studies in Epidemiology (STROBE) guidelines for cohort studies.

### Participants

Adults (≥19yrs) who were admitted to hospital between 17th January 2020 and 28^th^ February 2022 with confirmed or highly suspected SARS-CoV-2 infection leading to COVID-19 were included in this analysis: during the first wave, highly suspected cases were also eligible for inclusion, because SARS-CoV-2 was an emergent pathogen at that time and laboratory confirmation was dependent on availability of testing. SARS-CoV-2 infection was confirmed using reverse-transcriptase polymerase chain reaction (RT-PCR).

### Data sources

Data collected by research staff were entered into a standardised electronic case report form within a secure Research Electronic Data Capture (REDCap) database. Vaccination data were obtained from the national immunisation management system (NIMS), and deterministically linked to the ISARIC CCP-UK REDCap data using NHS number, which was collected as part of the ISARIC CCP-UK dataset.

Vaccination data were not available for Scottish patients and these analyses are therefore restricted to only English and Welsh ISARIC CCP-UK participants with a valid, linkable NHS number. Participants with invalid NHS numbers were excluded from the analysis after December 2020 to allow accurate linkage with vaccination data. Readmissions and those with erroneously recorded admission dates (eg, those with admission dates outside the scope of the study) were also removed prior to analysis.

### Variables

#### Immunocompromise

We considered patients to be immunocompromised if they met any of the following clinical criteria: solid organ transplant, active cancer diagnosis and treatment, congenital immune deficiency, human immunodeficiency (HIV) infection, in receipt of pre-admission immune-suppressive treatments, or pre-admission oral or intravenous steroids (see online supplement). For the purposes of analysis, these patients were only considered to be in one category, with the underlying reason for being immunocompromised being categorised by the following hierarchy: inherited immunological or metabolic disorder > solid organ transplant > cancer > HIV > pre-admission immunosuppressants > pre-admission steroids. Overlap between immunocompromising factors were visualised as part of the analysis.

#### Participant characteristics

patient demographics including age, sex, comorbidities, and ethnicity were recorded at hospital admission (see online supplement). Deprivation index was calculated using hospital level geographical data provided by the Office of National Statistics (ONS). Physiological parameters of the 4C Mortality Score (18) were used as markers of illness severity at presentation: respiratory rate (breaths per min), Glasgow Coma Scale, oxygen saturation (%), blood urea (mmol/L), blood C-reactive protein (CRP, mg/L). Symptoms recorded on admission are listed in the online supplement. If no symptom criteria were met, a patient was considered asymptomatic and was excluded from the analysis. If a patient met at least one of the criteria, they were considered to be symptomatic and were included.

In-hospital interventions were recorded including: critical care admission, level of respiratory support and treatments for COVID-19 including corticosteroids and interleukin-6 receptor blockers. For steroid and anti-IL-6 treatments, analyses were restricted to patients on oxygen therapy. Treatment with IL-6 receptor blockers was indicated by national guidance for patients with a C-reactive protein (CRP) blood level of 75mg/l or greater, or on respiratory support. Due to the nature of the data collection, pre-admission steroids were assessed through a free text search of pre-admission medication; dexamethasone, beclometasone, prednisolone, cortisone/hydrocortisone or betamethasone were included.

#### Vaccination

to investigate the effect of vaccination, we considered patients having received no vaccine doses or within 20 days of the first vaccine dose as being unprotected and therefore having no immunity to SARS-CoV-2; we then stratified patients as having received 1, 2, 3 and 4 or more doses, provided three weeks or more had elapsed between the first vaccination and symptom onset or positive RT-PCR test (whichever was earlier), or one week for subsequent doses, to allow immunity to develop (19).

#### Pandemic waves and SARS-CoV-2 variants

wave 1 was considered to be between 17^th^ January 2020, the date that the study protocol was activated in the UK, and 31^st^ August 2020, the nadir of hospital inpatient numbers between the first and second waves. This wave was largely accounted for by the B.1 lineage SARS-CoV-2 ancestral strain containing the D614G mutation. Wave 2 was defined as the period from 1^st^ September 2020 to 31^st^ March 2021, and comprised a mixture largely of B.1 D614G lineages and the alpha variant (B.1.1.7). The third wave, during which the alpha variant was replaced by the delta variant (B.1.617.2), was between 1^st^ April 2021 and 12^th^ December 2021. From 13^th^ December 2021 onwards, omicron (B.1.1.529) became the dominant circulating variant in the UK and out-competed delta (20). We refer to this period as the fourth wave.

#### Outcomes

the primary outcome was in-hospital mortality. Secondary outcomes were the use of oxygen, non-invasive ventilation, invasive mechanical ventilation and admission to critical care.

### Missing data

Due to the nature of such a large-scale, observational study, conducted during pandemic surge conditions, high degrees of missingness exist in multiple variables, particularly in the most recent wave dominated by the Omicron variant where recruitment has waned over time. As this study is predominantly descriptive, we have not performed any imputation to correct any data missingness, and instead are providing descriptions of the data as it is known.

### Statistical analyses

Continuous data are summarised as median (interquartile range) and categorical data as frequency (percentage). We employed statistical disclosure control (SDC) measures to protect patient confidentiality and anonymity.

To examine in-hospital mortality, we initially performed multivariable logistic regression adjusted for age, sex, number of comorbidities, ethnicity, deprivation index and vaccination (1). These variables were selected for inclusion in models based on their important effects previously described by ourselves and others (1, 21). Other outcome measures were the use of oxygen, non-invasive ventilation, intensive care admission and invasive ventilation, and were adjusted for the same variables. We subsequently explored whether changes in in-hospital mortality over time were different between immunocompetent and immunocompromised patients. A Bayesian logistic regression model was specified with weakly informative priors on model coefficients (four chains, 500 warmup, 2,000 iterations). The probability of death was determined for immunocompromised and immunocompetent patients in the first wave and the fourth wave, accounting for age, sex, socioeconomic deprivation, and comorbidity count. We then calculated the absolute risk difference for mortality in immunocompromised and immunocompetent patients in the first wave and compared this with the absolute risk difference in the omicron wave. We set comorbidity count to “2+”, and deprivation score to “2”, the most common level for each variable, and stratified these results by sex, age category, and vaccination status. All analysis was performed using the statistical software package R version 4.1.1 including the use of the associated packages tidyverse, finalfit and brms.

## Results

Between 17^th^ January 2020 and 28^th^ February 2022, data for 304,628 admissions of all ages were collected. Outcome data were available for 156,552 unique adult index admissions with symptomatic COVID-19 and valid NHS numbers (Figure S1). 134,598 (86%) patients were classified as immunocompetent, and 21,954 (14%) were immunocompromised (Figure S1). There was overlap between the immunocompromising conditions (Figure 1A), for example a large proportion of patients with previous solid organ transplant were also taking immunosuppressive medication. Using the hierarchical categorisation defined above, most patients that were identified as immunocompromised were either those taking immunosuppressive medication with no other documented immunocompromise (n=12,701 of 21,954, 58%), or those who had received recent cancer treatment (n=5,116, 23%). The numbers of patients with other conditions were: congenital immune deficiency (n=526, 2%), previous solid organ transplant (1,559, 7%), HIV/AIDS (498, 2%) and pre-admission steroids (n=1,554, 7%).

**Figure 1.**
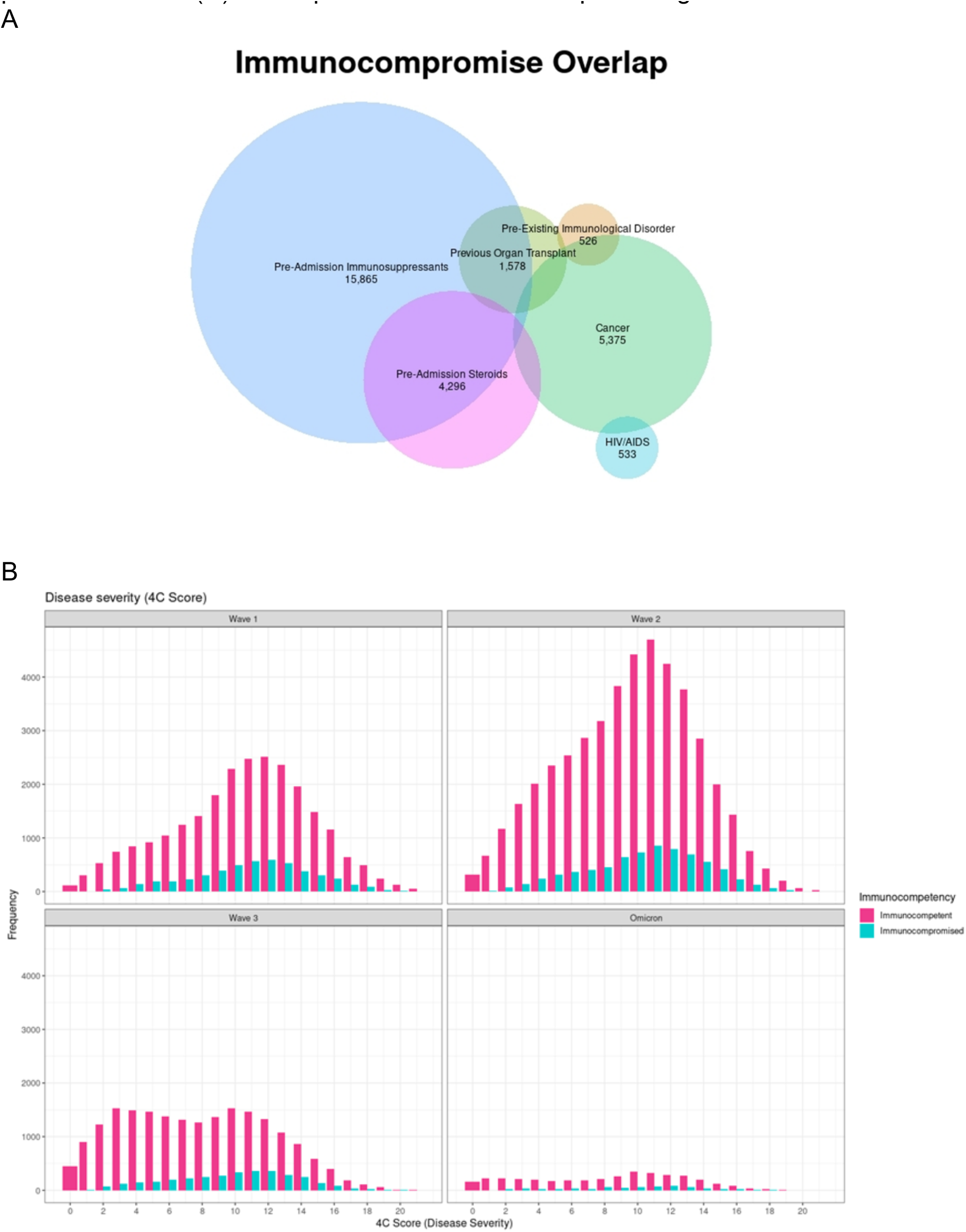
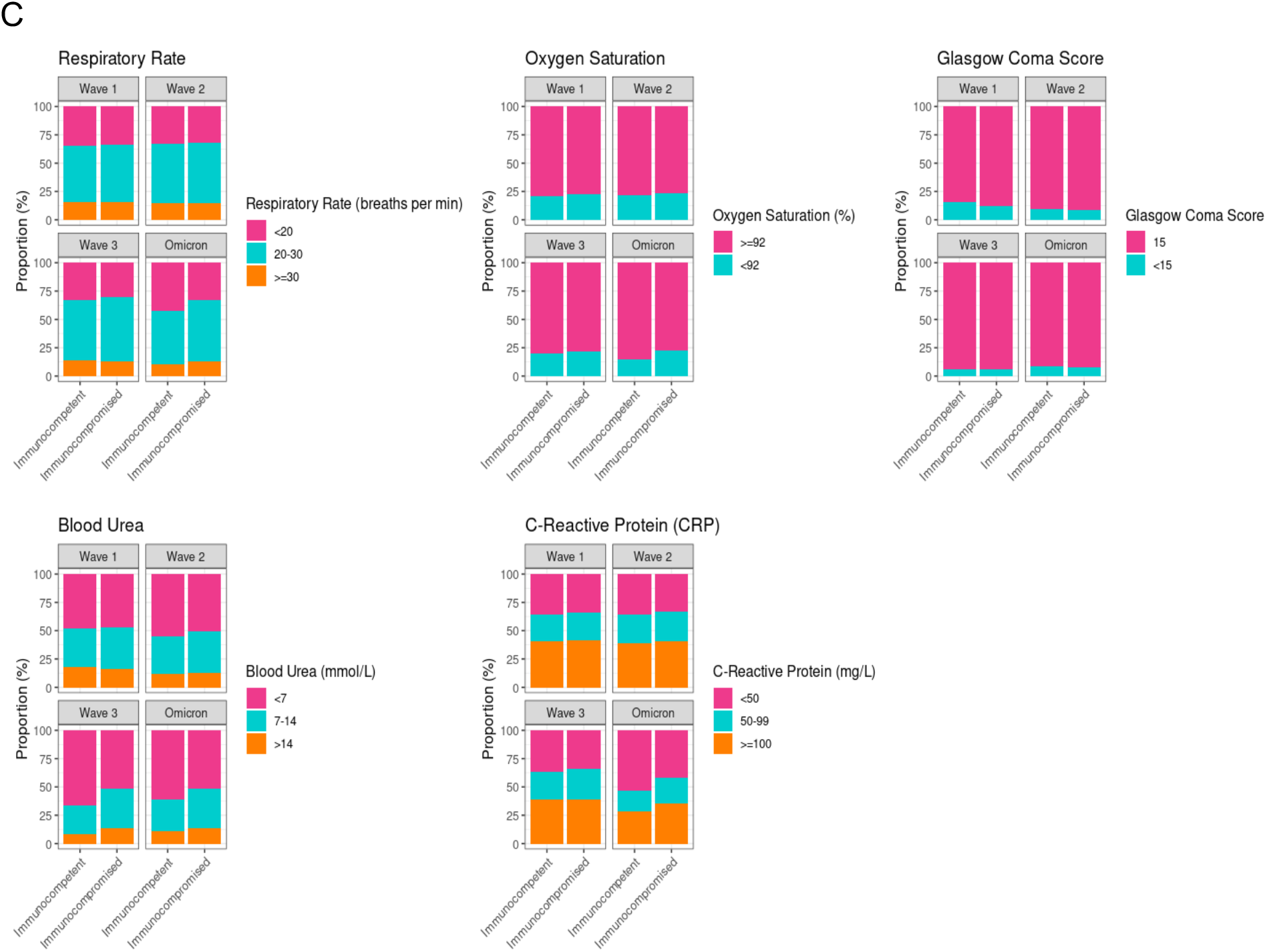
Illness severity and characteristics of the patients. (A) Illness severity over the course of the pandemic, measured using the 4C Mortality Score by immune status and pandemic wave. Pink bars: immunocompetent, green bars: immunocompromised. (B) Physiological components of the 4C Mortality Score stratified by immune status and pandemic wave. (C) Overlap between immunocompromising conditions

The median age of the immunocompetent patients was 69.5 years (IQR 53.4 to 82.0), and was slightly younger than that of the immunocompromised group which was 71.5 (IQR 58.7 to 80.3) (Table 1). Ages of immunocompromised patients varied depending on the aetiology of their immunocompromise. Those with cancer (median age 72.0. IQR (62.0 to 79.7)) and those on pre-admission immunosuppressants (median 72.5, IQR (59.1 to 81.2)) or corticosteroids (75.1 (63.0 to 83.6)) were considerably older than those with congenital immune deficiency (median 61.8, IQR (45.3 to 77.2)), solid organ transplant (median 62.4 (IQR 52.1 to 72.0)) and HIV (57.8 (IQR 49.8 to 70.5)). 95% (n=20,801 of 21,954) of patients with immunocompromise had at least one additional comorbidity (in addition to the reason for immunocompromise). Chronic pulmonary disease (n=6,190 of 21,954, (28%), immunocompromised) vs (n=18,495 of 134,598, (14%), immunocompetent), haematologic disease (n=2,041 (9%)) vs (n=3,665 (2%)), rheumatologic disease (n=4,540 (21%)) vs (n=12,901 (10%)), kidney disease (n=4,266 (19%)) vs (n=18,901 (14%)), and malignant neoplasm (n=5,113 (23%)) vs (n=8,484 (6%)) were more common in immunocompromised vs immunocompetent patients. Diabetes (n=6,042 (28%) immunocompromised) vs (n=36,322 (27%) immunocompetent), obesity (n=2,945 (13%)) vs (n=17,462 (13%)), hypertension (n=9,758 (44%)) vs (n=57,117 (42%)), chronic cardiac disease (n=6,607 (30%)) vs (n=35,597 (26%)) were similar in both groups (Table 1).

**Table 1.**
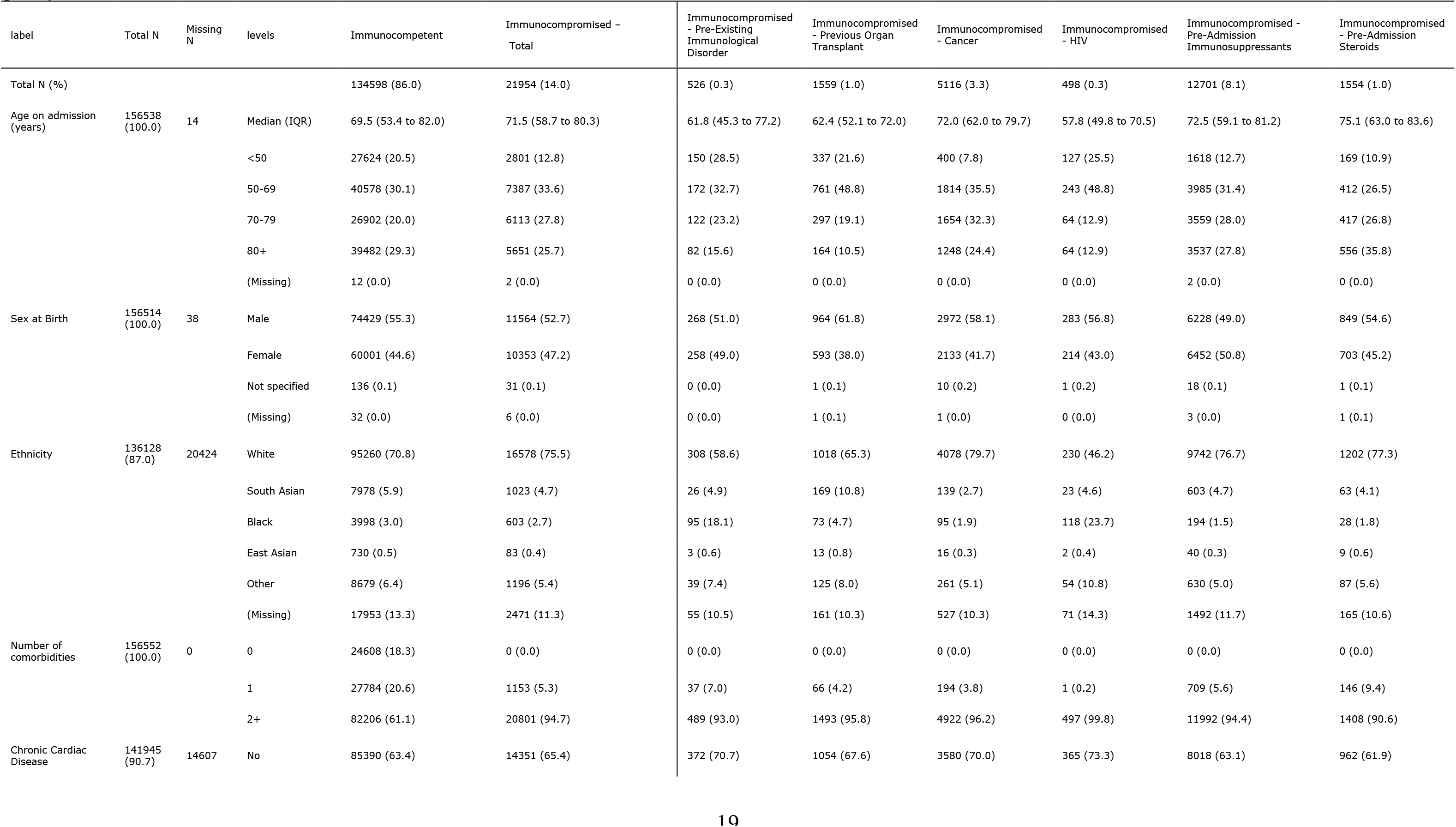
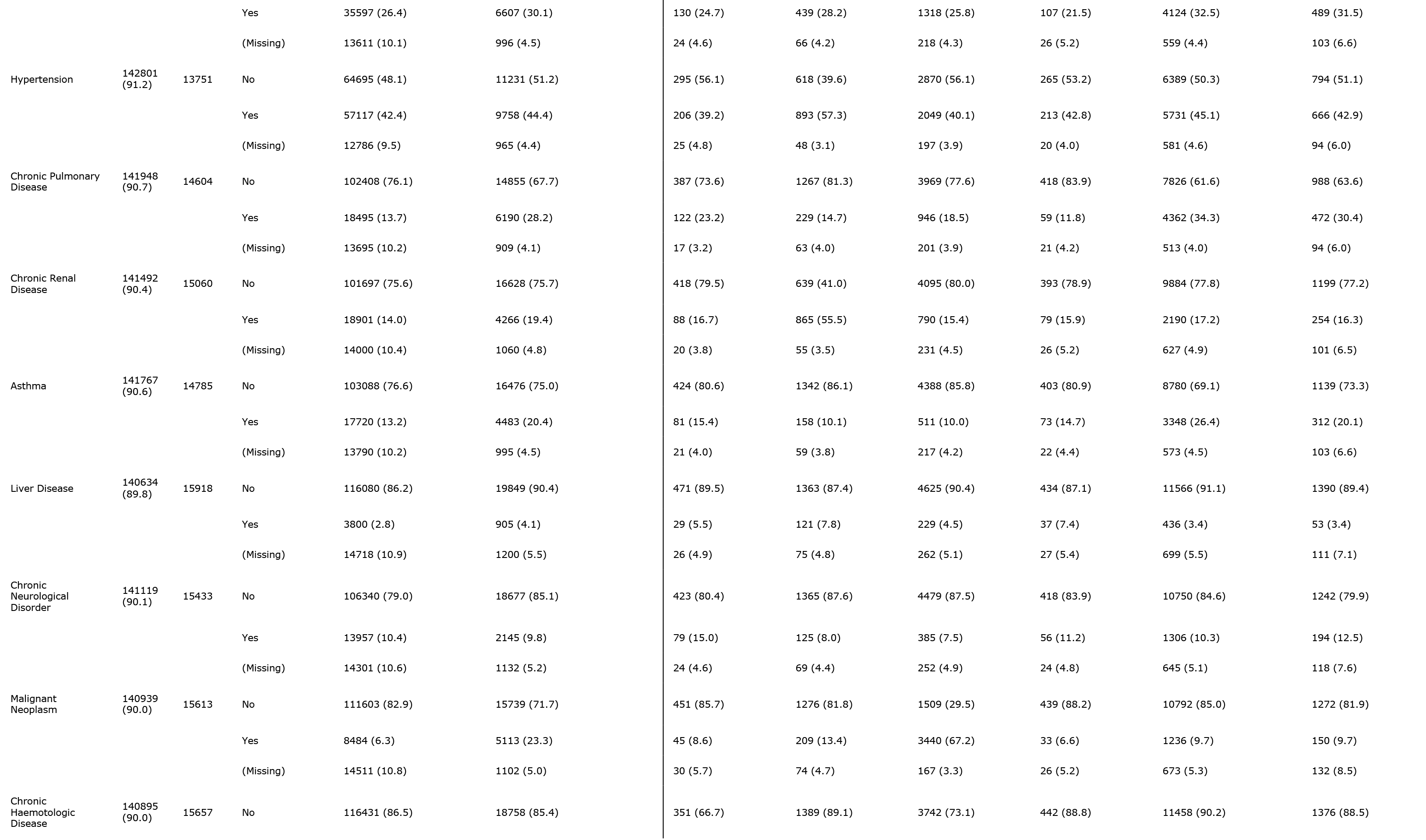
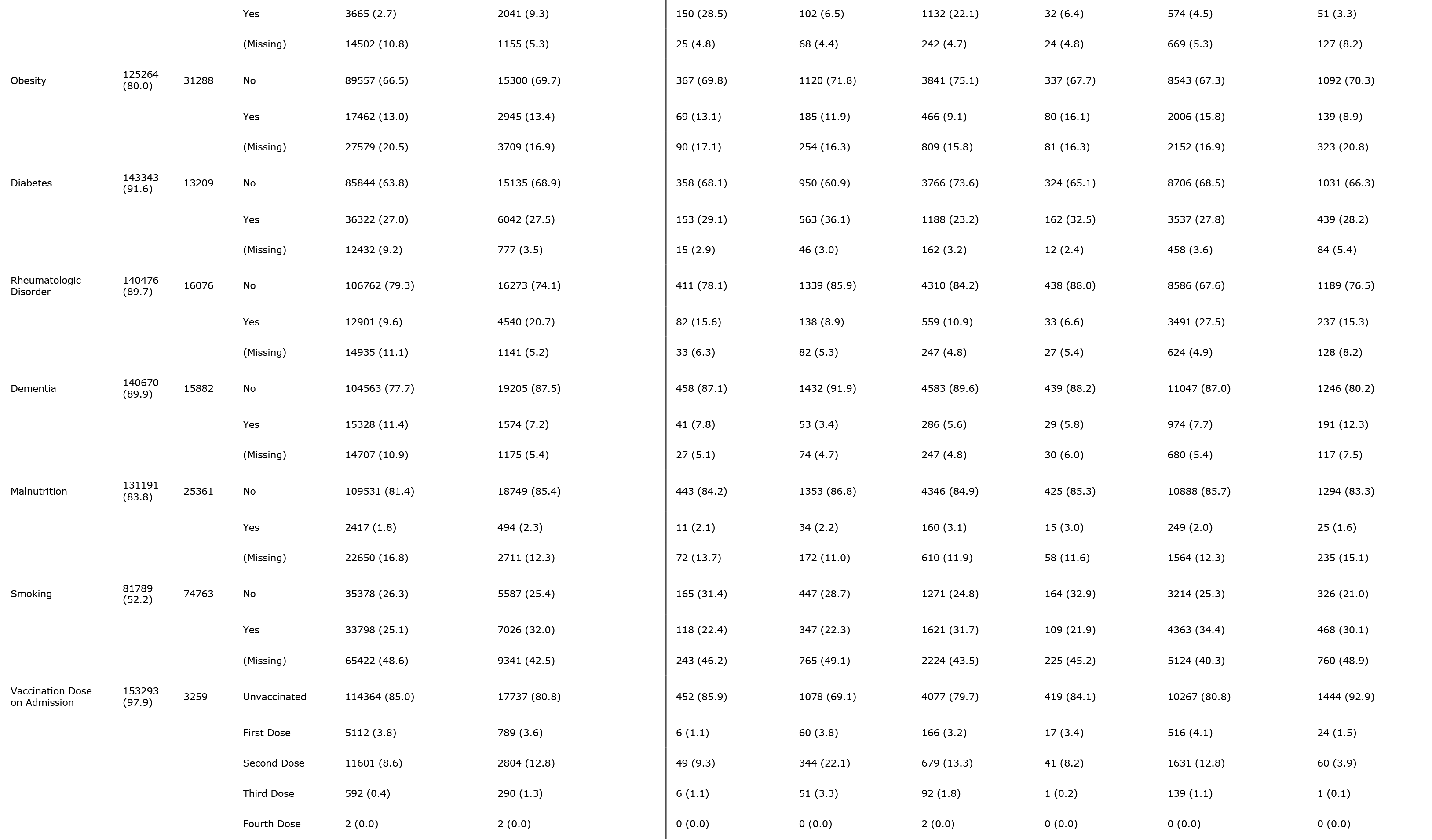
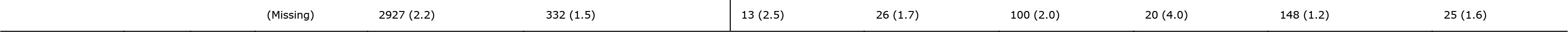
Demographics and comorbidities for immunocompetent and immunocompromised patients in the ISARIC WHO CCP-UK study. The immunocompromised patients are presented as both the, incorporating all conditions leading to immunocompromise, and by each group defined on the case record form.

Overall, 85% (n=114,364 of 134,598) of immunocompetent and 81% (n=17,737of 21,954) of immunocompromised patients hospitalised with COVID-19 were unvaccinated – 49% (n=76,948 of 156,552) were admitted before vaccines were available in waves 1 and 2 (Table 1, Figure S1). 5,112 (4%) of immunocompetent and 789 (4%) of immunocompromised patients admitted had received one dose of vaccine > 20 days earlier at the time of hospital admission. 11,601 (9%) immunocompetent and 2804 (13%) immunocompromised patients had received two doses > 7 days earlier. 592 (<1%) immunocompetent and 290 (1%) of immunocompromised patients had received 3 doses >7 days before the date of hospital admission (Table 1). Over time, the relative proportion of immunocompromised patients who were unvaccinated was less than for immunocompetent patients (Figure S2).

### Presenting symptoms and severity of illness

Illness severity at presentation to hospital as measured by the physiological components of the 4C Mortality Score was similar in the two groups at the beginning of the pandemic, suggesting no difference in the threshold for admission for immunocompromised patients (Figure 1B). Illness severity reduced in both groups over the course of the pandemic, although to a lesser extent in the immunocompromised group (Figure 1B and C). When compared with immunocompetent patients, in the omicron wave immunocompromised patients had higher respiratory rates (≥30bpm 67% vs immmunocompetent 57%), lower SaO2 (<92% 22% vs 15%), higher urea (urea ≥7mmol/l 49% vs 39%) and a higher inflammatory response (CRP ≥50mg/l 57% vs 47%) (Table S2, Figure 1C).

### Treatments received

When restricted to patients receiving oxygen, more patients in the immunocompromised group received corticosteroids than the immunocompetent group (Table S3, Figure S3). For patients on oxygen with a CRP>75, more immunocompetent patients received tocilizumab compared with immunocompromised patients (Table S3, Figure S4).

### Outcomes

Similar proportions of immunocompromised and immunocompetent patients received oxygen therapy and were admitted to critical care. Immunocompromised patients were more likely to receive non-invasive ventilation, and/or invasive mechanical ventilation (Table 2). 6,499 (29%) immunocompromised patients died, compared with 28,608 (21%) immunocompetent patients. The highest mortality rates were seen in patients with active cancer (n=1,818 (37%)) and those on pre-admission steroids (n=517 (34%)) (Table S4).

**Table 2.** Outcomes stratified by immune status and pandemic wave.

| label | levels | Immunocompetent - Wave 1 | Immunocompromised - Wave 1 | Immunocompetent - Wave 2 | Immunocompromised - Wave 2 | Immunocompetent - Wave 3 | Immunocompromised - Wave 3 | Immunocompetent - Wave 4 | Immunocompromised - Wave 4 |
| --- | --- | --- | --- | --- | --- | --- | --- | --- | --- |
| Oxygen | No | 10974 (31.2) | 1910 (28.0) | 16398 (25.7) | 2227 (22.8) | 7745 (29.0) | 1029 (24.0) | 3659 (51.6) | 361 (40.1) |
|  | Yes | 24227 (68.8) | 4918 (72.0) | 47468 (74.3) | 7549 (77.2) | 18999 (71.0) | 3258 (76.0) | 3434 (48.4) | 540 (59.9) |
| Critical Care Admission | No | 30785 (86.9) | 5952 (86.9) | 52873 (82.5) | 8101 (82.7) | 22469 (83.9) | 3514 (82.0) | 6611 (92.4) | 823 (90.8) |
|  | Yes | 4637 (13.1) | 896 (13.1) | 11223 (17.5) | 1693 (17.3) | 4306 (16.1) | 773 (18.0) | 544 (7.6) | 83 (9.2) |
| Non-Invasive Ventilation | No | 29970 (85.4) | 5660 (83.2) | 50363 (79.4) | 7449 (76.8) | 20925 (78.9) | 3181 (74.7) | 6362 (90.1) | 762 (84.9) |
|  | Yes | 5106 (14.6) | 1145 (16.8) | 13035 (20.6) | 2252 (23.2) | 5608 (21.1) | 1078 (25.3) | 696 (9.9) | 136 (15.1) |
| Invasive Ventilation | No | 32524 (92.4) | 6384 (93.6) | 59058 (93.0) | 8973 (92.2) | 25074 (94.7) | 3948 (92.8) | 6875 (97.6) | 866 (96.7) |
|  | Yes | 2664 (7.6) | 435 (6.4) | 4416 (7.0) | 754 (7.8) | 1415 (5.3) | 308 (7.2) | 170 (2.4) | 30 (3.3) |
| Death | No | 24766 (70.2) | 4379 (64.3) | 48439 (77.4) | 6551 (68.6) | 23276 (87.9) | 3317 (78.3) | 5726 (88.7) | 662 (81.2) |
|  | Yes | 10495 (29.8) | 2430 (35.7) | 14177 (22.6) | 2998 (31.4) | 3210 (12.1) | 918 (21.7) | 726 (11.3) | 153 (18.8) |

After adjustment for age, sex, ethnicity, deprivation and comorbidities, the odds ratio (OR) for death in the immunocompromised group was 1.44 (95% CI 1.39-1.5, p < 0.001) (Figure 2A). The ORs of several other outcomes were significantly different for immunocompromised relative to immunocompetent patients, though with much smaller effect sizes. The OR for receiving oxygen was 1.10 (95% CI 1.06, 1.15, p<0.001), for critical care admission was 1.08 (95%CI 1.03, 1.13, p=0.001), receiving non-invasive ventilation 1.17 (95%CI 1.12, 1.22, p<0.001), and invasive ventilation 1.10 (95%CI 1.03, 1.18, p=0.005) (Figure 2A).

**Figure 2.**
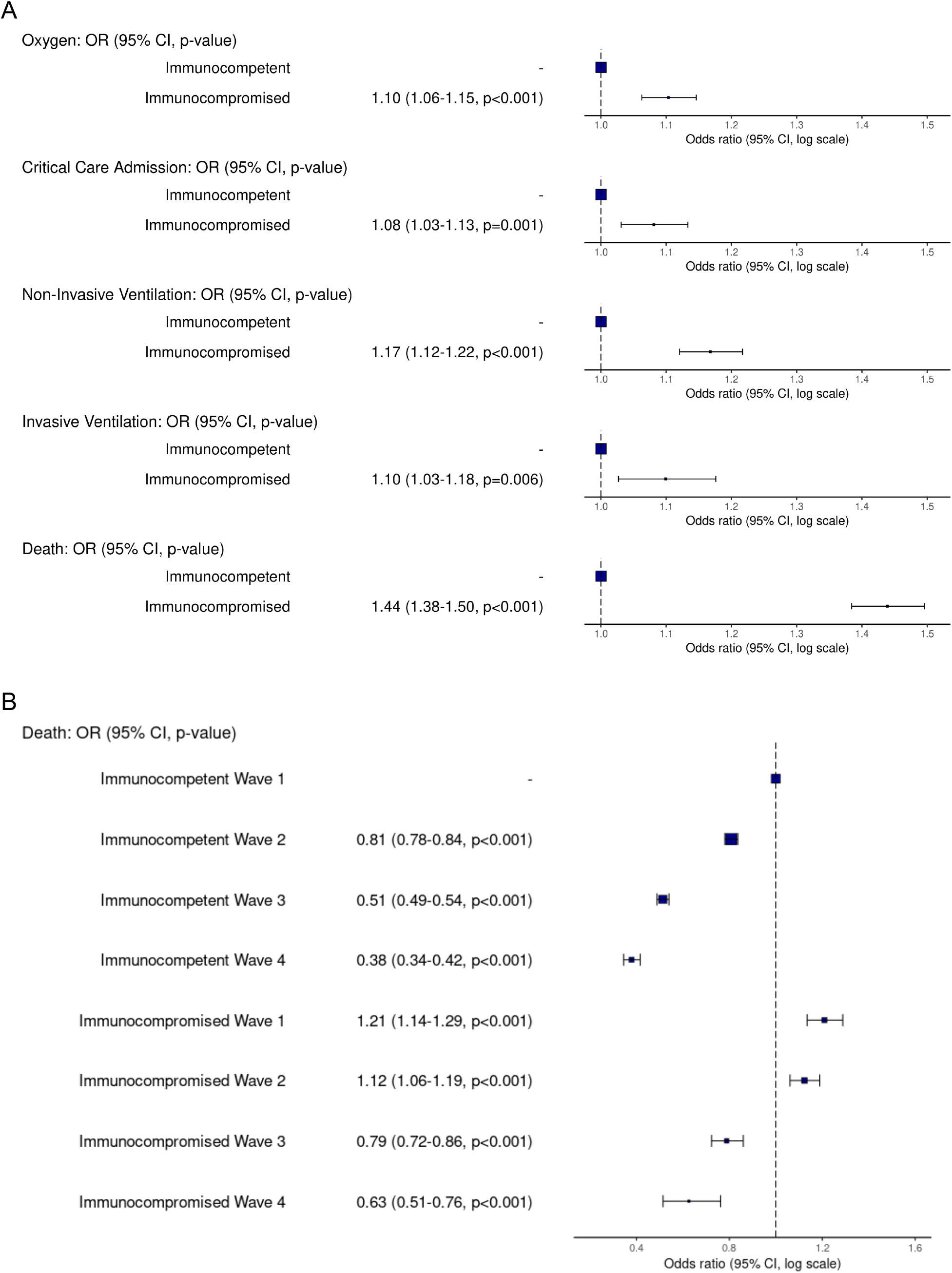
Outcomes of hospitalised immunocompromised patients, compared with immunocompetent patients. Odds ratios (ORs) from multivariable logistic regression and 95% confidence intervals (CI) for (A) outcomes of death, oxygen use, critical care admission, non-invasive and invasive ventilation, and (B) for death in the first 4 pandemic waves in the UK. ORs are adjusted for age, sex, ethnicity, deprivation, and comorbidities.

### Changes in mortality over time

Mortality for both immunocompetent and immunocompromised groups reduced over time. Crude in-hospital mortality of immunocompetent patients in the first wave was 30%, compared with 36% in immunocompromised patients. In the fourth (omicron) wave, the mortality was 11% for immunocompetent patients, and 19% for immunocompromised patients (Table 2).

After adjustment for age, sex, comorbidity count, deprivation and vaccination status, compared with immunocompetent patients in wave 1, the OR for mortality remained elevated in all waves in immunocompromised patients (Figure 2B). Univariable analysis showed the same trend (Figure S5). In the fourth wave, nearly a year after the vaccination program was initiated, the OR for death compared with the first wave was 0.38 (95%CI 0.34, 0.42, p<0.001) for immunocompetent patients and 0.63 (95%CI 0.51, 0.76, p<0.001) for immunocompromised patients (Figure 2B). There was a decline in the number of patients enrolled in the study over time (Figure S6).

To investigate this further, we calculated the difference in risk of death between the first and fourth waves for both immunocompetent and immunocompromised patients. using the Bayesian framework described in the methods, the probability that this difference was less for immunocompromised patients (indicating less improvement over the course of the pandemic) was always greater than 67% (Figure 3). Across all ages, and both sexes, in-hospital mortality for immunocompromised patients improved less than for immunocompetent patients. This was particularly evident with increasing age: the probability of the reduction in hospital mortality being less for immunocompromised patients aged 50-69yrs was 88% for men and 83% for women, and for those >80yrs was 99% for men, and 98% for women.

**Figure 3:**
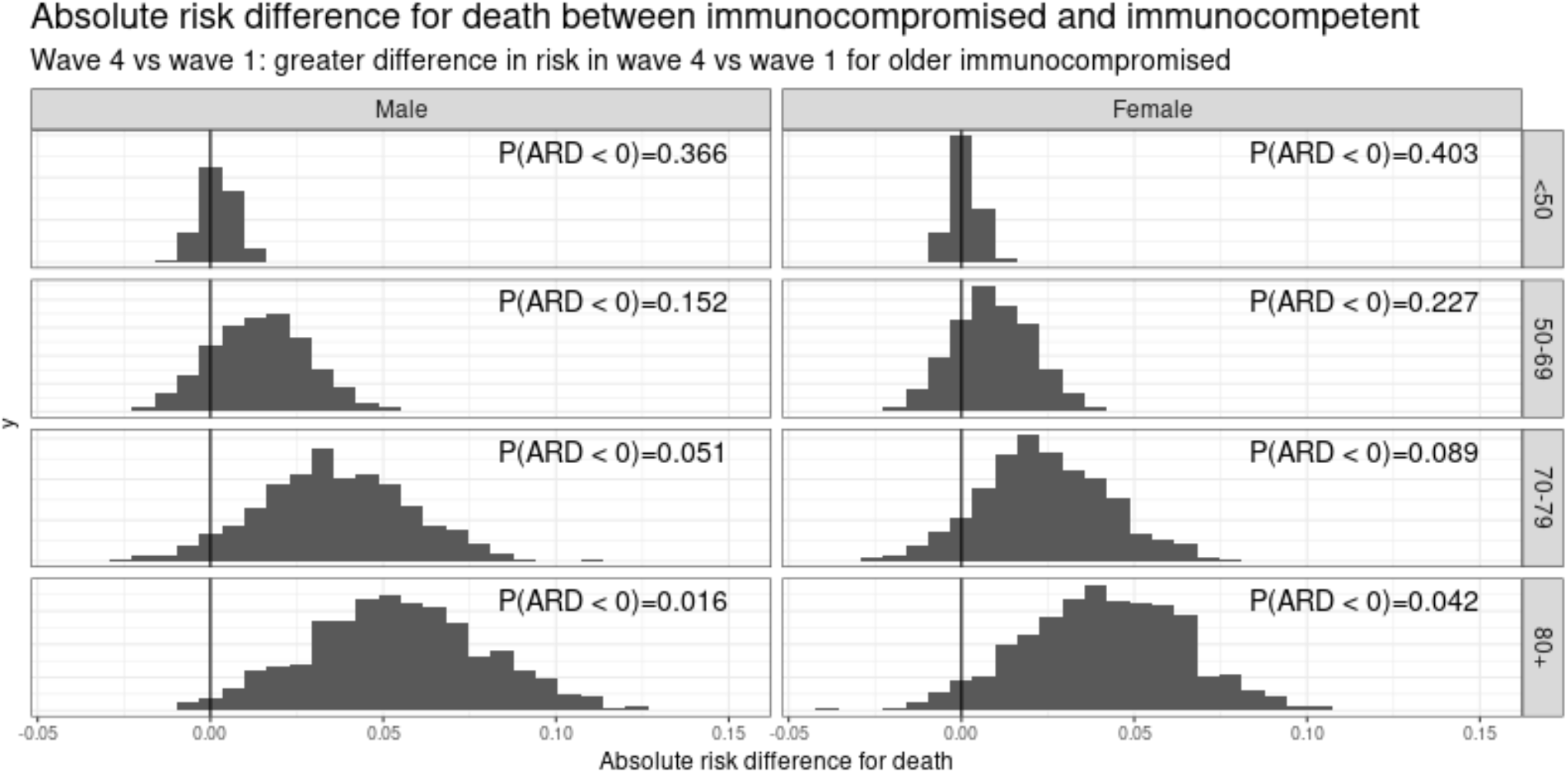
Absolute risk difference (ARD) for death between immunocompromised and immunocompetent. The difference in risk of death in the first wave, and the fourth wave, for immunocompetent and immunocompromised patients was modelled using Bayesian logistic regression, and the probability that the risk of death across the two waves reduced more in the immunocompetent that the immunocompromised was calculated. The analysis was stratified by age and sex.

## Discussion

Using the ISARIC CCP-UK cohort, we found that immunocompromised people admitted to hospital with COVID-19 had greater adjusted mortality than the general in-patient population. Mortality in this group has also not fallen to the same extent as the immunocompetent patients. Over the course of the pandemic, immunocompromised patients have seen less reduction in severity at presentation to hospital and less improvement in in-hospital mortality than immunocompetent patients.

Large population studies early in the pandemic have suggested that patients who are immunocompromised are more likely to die from COVID-19 than those who are immunocompetent (16). However, several studies, including one of the largest to date (22), have not shown an increased risk of in-hospital mortality in immunocompromised patients when compared to those with no known immunodeficiency. One possible explanation for this is that immunocompromised patients are more likely to be admitted to hospital, but once hospitalised, the outcomes are similar. Our data challenge this hypothesis. Once admitted, the odds of in-hospital mortality (adjusted for age, sex, ethnicity, vaccination and co-morbidities) were higher for the immunocompromised group, with less improvement in outcome over the course of the pandemic compared with the immunocompetent group. Disease severity (physiological derangement) at presentation was similar for the two groups early in the pandemic. Whilst severity at presentation has decreased for both groups over time, as the pandemic progressed, the improvement seen was greater for immunocompetent patients. Immunocompromised patients are now on average more ill relative to immunocompetent patients. Other possibilities for differences between studies potentially include selection of patient groups to include, criteria for hospital admission and socio-ethnodemographic variation.

Immunocompromised patients with COVID-19 received steroids more frequently during their admission than immunocompetent patients. This might indicate a continuation of pre-admission, non-covid related steroid treatment. The less frequent use of tocilizumab, on the other hand (in patients who met the criteria for use), may indicate concern about the net state of immune suppression and careful weighing of risk benefit in the minds of treating clinicians. To better inform the use of anti-inflammatory treatments in this group, future studies targeted at immunocompromised patients, or with recruitment stratified by immune status, would be needed.

There was a reduction over time in the proportion of patients admitted after vaccination compared with those unvaccinated. However, the risk difference between immunocompromised and immunocompetent patients widened between the first wave, and the fourth wave, by which time vaccination levels in the population were high. In combination with our earlier work (23), this indicates that this group of patients may remain more vulnerable than the general population even after vaccination. Selection of SARS-CoV-2 variants is largely a function of transmissibility (24, 25), or immune escape (26), and a future variant could still have higher intrinsic virulence than omicron. In this case, immunocompromised patients might be more vulnerable despite vaccination.

Our findings differ to those of Andersen *et al*. (22), which did not find an increase in mortality in immunocompromised patients from 42 US health systems. A difference in age may explain this discrepancy. The mean age of patients reported by Andersen *et al*. (59 years) was younger than our median age (71.5 years), suggesting that the difference between the two studies could be that here we are describing an older, more co-morbid population. In our study chronic pulmonary, haematologic, rheumatologic, kidney and liver disease were all more common in immunocompromised patients, which may reflect either the reasons for immunosuppression, or complications thereof. However, comorbidities such as heart disease and hypertension were not more common in the immunocompromised patients, indicating that differences in comorbidity were not the only reason for the difference in outcomes between the groups.

The ISARIC CCP-UK cohort is one of the largest datasets of hospitalised patients with COVID-19 in the world and has enabled us to study small subgroups such as those with congenital immune deficiencies, which have not been captured in smaller cohorts. The ISARIC CCP-UK dataset has detailed information regarding severity of illness at presentation which allowed us to explore whether the threshold for hospital admission differed between patient groups. We were also able to follow patient journeys through the hospital looking at in-hospital interventions and respiratory support which are lacking from routine healthcare datasets. There are some weaknesses to this study. We did not collect data on the detailed drug histories of patients prior to admission, and so we did not have information on which drug was responsible for immunocompromise. We lacked data on pre-admission steroid doses, meaning that we were unable to assess the extent to which steroid use contributed to immune compromise. Missing data in the medical and drug histories means that we may have underestimated the overall effect size by categorising some immunocompromised patients as immunocompetent.

We have observed that in hospital mortality for patients who are clinically extremely vulnerable with immunocompromise has fallen less than for than immunocompetent patients in the ISARIC CCP-UK dataset. Despite the benefits of vaccination, this group still lags behind the general patient population in the improvements in outcomes after hospitalisation. A number of interventions, such as remdesivir, molnupiravir and nirmatrelvir/ritonavir are now available to reduce the risk of progression to severe COVID-19 and hospitalisation in this patient group (27, 28). However, some immunocompromised patients do not respond well to COVID-19 vaccines (29). and several monoclonal antibody treatments have now ceased to be effective due to antigenic variation of the omicron variant (30). However, tixagevimab/cilgavimab retains activity against omicron sub-variants BA.2, BA.4 and BA.5, but is not available for routine clinical care in the UK. The use of tixagevimab/cilgavimab remains a policy option and may close the gap between immunocompromised patients and the general population.

## Data Availability

Data cannot be shared publicly because of they contain person identifiable information. ISARIC4C welcomes applications for data and material access through our Independent Data and Material Access Committee (https://isaric4c.net) for researchers who meet the criteria for access to confidential data.

## Acknowledgements

This work is supported by grants from: the National Institute for Health Research (NIHR) [award CO-CIN-01], the Medical Research Council [grant MC_PC_19059 and MR/V028979/1], the Chief Scientist Office, Scotland, and by the NIHR Health Protection Research Unit (HPRU) in Emerging and Zoonotic Infections at University of Liverpool in partnership with the UK Health Security Agency (UK-HSA), in collaboration with Liverpool School of Tropical Medicine and the University of Oxford [award 200907], NIHR HPRU in Respiratory Infections at Imperial College London with UK-HSA [award 200927], Wellcome Trust and Department for International Development [215091/Z/18/Z], the Bill and Melinda Gates Foundation [OPP1209135], Liverpool Experimental Cancer Medicine Centre (Grant Reference: C18616/A25153), NIHR Biomedical Research Centre at Imperial College London [IS-BRC-1215-20013], EU Platform foR European Preparedness Against (Re-) emerging Epidemics (PREPARE) [FP7 project 602525]. We acknowledge the NIHR Clinical Research Network for providing infrastructure support for this research. This research is part of the Data and Connectivity National Core Study, led by Health Data Research UK in partnership with the Office for National Statistics and funded by UK Research and Innovation (grant ref MC_PC_20029). LT is supported by a Wellcome Trust clinical career development fellowship a [205228/Z/16/Z]. PJMO is supported by a NIHR Senior Investigator Award [award 201385]. DGW is supported by an NIHR Advanced Fellowship [NIHR300669]. For the purpose of Open Access, the authors have applied a CC-BY public copyright licence to any Author Accepted Manuscript version arising from this submission. The views expressed are those of the authors and not necessarily those of the DHSC, DID, NIHR, MRC, Wellcome Trust or UK-HSA.

## Data availability

This work uses data provided by patients and collected by the NHS as part of their care and support #DataSavesLives. The CO-CIN data was collated by ISARIC4C Investigators. ISARIC4C welcomes applications for data and material access through our Independent Data and Material Access Committee (https://isaric4c.net).

## ISARIC4C investigators

*\*Co-Lead Investigator (Consortium Lead)*: J Kenneth Baillie.

*\*Co-Lead Investigator (Sponsor & Protocol CI)*: Malcolm G Semple.

*\*Co-Lead Investigator (Imperial College)*: Peter JM Openshaw.

### Co-Investigator

Beatrice Alex, Petros Andrikopoulos, Benjamin Bach, Wendy S Barclay, Debby Bogaert, Meera Chand, Kanta Chechi, Graham S Cooke, Ana da Silva Filipe, Thushan de Silva, Annemarie B Docherty, Gonçalo dos Santos Correia, Marc-Emmanuel Dumas, Jake Dunning, Tom Fletcher, Christoper A Green, William Greenhalf, Julian L Griffin, Rishi K Gupta, Ewen M Harrison, Julian A Hiscox, Antonia Ying Wai Ho, Karl Holden, Peter W Horby, Samreen Ijaz, Saye Khoo, Paul Klenerman, Andrew Law, Matthew R Lewis, Sonia Liggi, Wei Shen Lim, Lynn Maslen, Alexander J Mentzer, Laura Merson, Alison M Meynert, Shona C Moore, Mahdad Noursadeghi, Michael Olanipekun, Anthonia Osagie, Massimo Palmarini, Carlo Palmieri, William A Paxton, Georgios Pollakis, Nicholas Price, Andrew Rambaut, David L Robertson, Clark D Russell, Vanessa Sancho-Shimizu, Caroline J Sands, Janet T Scott, Louise Sigfrid, Tom Solomon, Shiranee Sriskandan, David Stuart, Charlotte Summers, Olivia V Swann, Zoltan Takats, Panteleimon Takis, Richard S Tedder, AA Roger Thompson, Emma C Thomson, Ryan S Thwaites, Lance CW Turtle, Maria Zambon.

### Data Analyst

Thomas M Drake, Cameron J Fairfield, Stephen R Knight, Kenneth A Mclean, Derek Murphy, Lisa Norman, Riinu Pius, Catherine A Shaw.

### Data and Information System Manager

Marie Connor, Jo Dalton, Carrol Gamble, Michelle Girvan, Sophie Halpin, Janet Harrison, Clare Jackson, James Lee, Laura Marsh, Daniel Plotkin, Stephanie Roberts, Egle Saviciute.

### Data Integration and Presentation

Sara Clohisey, Ross Hendry, Susan Knight, Eva Lahnsteiner, Andrew Law, Gary Leeming, Lucy Norris, James Scott-Brown, Sarah Tait, Murray Wham.

### ISARIC Clinical Coordinator

Gail Carson.

### Edinburgh Laboratory Staff and Volunteers

Richard Clark, Audrey Coutts, Lorna Donnelly, Angie Fawkes, Tammy Gilchrist, Katarzyna Hafezi, Louise MacGillivray, Alan Maclean, Sarah McCafferty, Kirstie Morrice, Lee Murphy, Nicola Wrobel.

### Local Principal Investigators

Kayode Adeniji, Daniel Agranoff, Ken Agwuh, Dhiraj Ail, Erin L. Aldera, Ana Alegria, Sam Allen, Brian Angus, Abdul Ashish, Dougal Atkinson, Shahedal Bari, Gavin Barlow, Stella Barnass, Nicholas Barrett, Christopher Bassford, Sneha Basude, David Baxter, Michael Beadsworth, Jolanta Bernatoniene, John Berridge, Colin Berry, Nicola Best, Pieter Bothma, Robin Brittain-Long, Naomi Bulteel, Tom Burden, Andrew Burtenshaw, Vikki Caruth, David Chadwick, David Chadwick, Duncan Chambler, Nigel Chee, Jenny Child, Srikanth Chukkambotla, Tom Clark, Paul Collini, Catherine Cosgrove, Jason Cupitt, Maria-Teresa Cutino-Moguel, Paul Dark, Chris Dawson, Samir Dervisevic, Phil Donnison, Sam Douthwaite, Andrew Drummond, Ingrid DuRand, Ahilanadan Dushianthan, Tristan Dyer, Cariad Evans, Chi Eziefula, Chrisopher Fegan, Adam Finn, Duncan Fullerton, Sanjeev Garg, Sanjeev Garg, Atul Garg, Effrossyni Gkrania-Klotsas, Jo Godden, Arthur Goldsmith, Clive Graham, Tassos Grammatikopoulos, Elaine Hardy, Stuart Hartshorn, Daniel Harvey, Peter Havalda, Daniel B Hawcutt, Maria Hobrok, Luke Hodgson, Anil Hormis, Joanne Howard, Michael Jacobs, Susan Jain, Paul Jennings, Agilan Kaliappan, Vidya Kasipandian, Stephen Kegg, Michael Kelsey, Jason Kendall, Caroline Kerrison, Ian Kerslake, Oliver Koch, Gouri Koduri, George Koshy, Shondipon Laha, Steven Laird, Susan Larkin, Tamas Leiner, Patrick Lillie, James Limb, Vanessa Linnett, Jeff Little, Mark Lyttle, Michael MacMahon, Emily MacNaughton, Ravish Mankregod, Huw Masson, Elijah Matovu, Katherine McCullough, Ruth McEwen, Manjula Meda, Gary Mills, Jane Minton, Mariyam Mirfenderesky, Kavya Mohandas, Quen Mok, James Moon, Elinoor Moore, Patrick Morgan, Craig Morris, Katherine Mortimore, Samuel Moses, Mbiye Mpenge, Rohinton Mulla, Michael Murphy, Thapas Nagarajan, Megan Nagel, Mark Nelson, Lillian Norris, Matthew K. O’Shea, Marlies Ostermann, Igor Otahal, Mark Pais, Carlo Palmieri, Selva Panchatsharam, Danai Papakonstantinou, Padmasayee Papineni, Hassan Paraiso, Brij Patel, Natalie Pattison, Justin Pepperell, Mark Peters, Mandeep Phull, Stefania Pintus, Tim Planche, Frank Post, David Price, Rachel Prout, Nikolas Rae, Henrik Reschreiter, Tim Reynolds, Neil Richardson, Mark Roberts, Devender Roberts, Alistair Rose, Guy Rousseau, Bobby Ruge, Brendan Ryan, Taranprit Saluja, Sarah Cole, Matthias L Schmid, Aarti Shah, Manu Shankar-Hari, Prad Shanmuga, Anil Sharma, Anna Shawcross, Jagtur Singh Pooni, Jeremy Sizer, Richard Smith, Catherine Snelson, Nick Spittle, Nikki Staines, Tom Stambach, Richard Stewart, Pradeep Subudhi, Tamas Szakmany, Kate Tatham, Jo Thomas, Chris Thompson, Robert Thompson, Ascanio Tridente, Darell Tupper-Carey, Mary Twagira, Nick Vallotton, Rama Vancheeswaran, Lisa Vincent-Smith, Shico Visuvanathan, Alan Vuylsteke, Sam Waddy, Rachel Wake, Andrew Walden, Ingeborg Welters, Tony Whitehouse, Paul Whittaker, Ashley Whittington, Meme Wijesinghe, Martin Williams, Lawrence Wilson, Stephen Winchester, Martin Wiselka, Adam Wolverson, Daniel G Wootton, Andrew Workman, Bryan Yates, Peter Young. *Material Management*: Sarah E McDonald, Victoria Shaw.

### Outbreak Laboratory Staff and Volunteers

Katie A. Ahmed, Jane A Armstrong, Milton Ashworth, Innocent G Asiimwe, Siddharth Bakshi, Samantha L Barlow, Laura Booth, Benjamin Brennan, Katie Bullock, Nicola Carlucci, Emily Cass, Benjamin WA Catterall, Jordan J Clark, Emily A Clarke, Sarah Cole, Louise Cooper, Helen Cox, Christopher Davis, Oslem Dincarslan, Alejandra Doce Carracedo, Chris Dunn, Philip Dyer, Angela Elliott, Anthony Evans, Lorna Finch, Lewis WS Fisher, Lisa Flaherty, Terry Foster, Isabel Garcia-Dorival, Philip Gunning, Catherine Hartley, Anthony Holmes, Rebecca L Jensen, Christopher B Jones, Trevor R Jones, Shadia Khandaker, Katharine King, Robyn T. Kiy, Chrysa Koukorava, Annette Lake, Suzannah Lant, Diane Latawiec, Lara Lavelle-Langham, Daniella Lefteri, Lauren Lett, Lucia A Livoti, Maria Mancini, Hannah Massey, Nicole Maziere, Sarah McDonald, Laurence McEvoy, John McLauchlan, Soeren Metelmann, Nahida S Miah, Joanna Middleton, Joyce Mitchell, Shona C Moore, Ellen G Murphy, Rebekah Penrice-Randal, Jack Pilgrim, Tessa Prince, Will Reynolds, P. Matthew Ridley, Debby Sales, Victoria E Shaw, Rebecca K Shears, Benjamin Small, Krishanthi S Subramaniam, Agnieska Szemiel, Aislynn Taggart, Jolanta Tanianis-Hughes, Jordan Thomas, Erwan Trochu, Libby van Tonder, Eve Wilcock, J. Eunice Zhang.

### Patient Engagement

Seán Keating.

### Project Administrator

Cara Donegan, Rebecca G. Spencer, Lauren Obosi. *Project Manager*: Chloe Donohue, Fiona Griffiths, Hayley Hardwick, Wilna Oosthuyzen.

## Supplementary data

**Table S1.** Comorbidities in immunocompetent versus immunocompromised patients stratified by pandemic wave.

| label | levels | Immunocompetent - Wave 1 | Immunocompromised - Wave 1 | Immunocompetent - Wave 2 | Immunocompromised - Wave 2 | Immunocompetent - Wave 3 | Immunocompromised - Wave 3 | Immunocompetent - Wave 4 | Immunocompromised - Wave 4 |
| --- | --- | --- | --- | --- | --- | --- | --- | --- | --- |
| Age on admission (years) | Median (IQR) | 75.3 (59.7 to 84.9) | 74.1 (62.0 to 82.5) | 70.2 (55.6 to 81.9) | 71.3 (59.0 to 80.1) | 58.7 (42.2 to 75.2) | 66.9 (53.9 to 76.6) | 66.8 (45.5 to 81.5) | 69.2 (56.2 to 78.6) |
|  | <50 | 4716 (13.3) | 641 (9.3) | 10867 (16.8) | 1209 (12.3) | 9867 (36.4) | 806 (18.6) | 2174 (29.4) | 145 (15.7) |
|  | 50-69 | 9402 (26.5) | 2010 (29.3) | 21188 (32.8) | 3389 (34.4) | 8129 (30.0) | 1658 (38.3) | 1859 (25.2) | 330 (35.8) |
|  | 70-79 | 7472 (21.0) | 1977 (28.8) | 13751 (21.3) | 2764 (28.1) | 4381 (16.2) | 1110 (25.7) | 1298 (17.6) | 262 (28.4) |
|  | 80+ | 13931 (39.2) | 2230 (32.5) | 18755 (29.1) | 2485 (25.2) | 4738 (17.5) | 751 (17.4) | 2058 (27.9) | 185 (20.1) |
| Sex at Birth | Male | 19501 (54.9) | 3684 (53.7) | 36124 (56.0) | 5196 (52.8) | 15137 (55.8) | 2234 (51.7) | 3667 (49.6) | 450 (48.8) |
|  | Female | 15976 (45.0) | 3163 (46.1) | 28371 (43.9) | 4636 (47.1) | 11948 (44.1) | 2083 (48.2) | 3706 (50.2) | 471 (51.1) |
|  | Not specified | 41 (0.1) | 8 (0.1) | 60 (0.1) | 15 (0.2) | 22 (0.1) | 7 (0.2) | 13 (0.2) | 1 (0.1) |
| Ethnicity | White | 25322 (82.0) | 5207 (85.0) | 46492 (82.5) | 7399 (85.0) | 18501 (79.8) | 3262 (85.0) | 4945 (79.7) | 710 (86.8) |
|  | South Asian | 1856 (6.0) | 300 (4.9) | 4094 (7.3) | 490 (5.6) | 1610 (6.9) | 201 (5.2) | 418 (6.7) | 32 (3.9) |
|  | Black | 1159 (3.8) | 220 (3.6) | 1542 (2.7) | 229 (2.6) | 999 (4.3) | 127 (3.3) | 298 (4.8) | 27 (3.3) |
|  | East Asian | 243 (0.8) | 30 (0.5) | 307 (0.5) | 38 (0.4) | 158 (0.7) | 14 (0.4) | 22 (0.4) | 1 (0.1) |
|  | Other | 2313 (7.5) | 368 (6.0) | 3920 (7.0) | 547 (6.3) | 1924 (8.3) | 233 (6.1) | 522 (8.4) | 48 (5.9) |
| Number of comorbidities | 0 | 4378 (12.3) | 0 (0.0) | 10230 (15.8) | 0 (0.0) | 8238 (30.4) | 0 (0.0) | 1762 (23.8) | 0 (0.0) |
|  | 1 | 6868 (19.3) | 285 (4.2) | 13649 (21.1) | 489 (5.0) | 5809 (21.4) | 335 (7.7) | 1458 (19.7) | 44 (4.8) |
|  | 2+ | 24279 (68.3) | 6573 (95.8) | 40688 (63.0) | 9359 (95.0) | 13070 (48.2) | 3991 (92.3) | 4169 (56.4) | 878 (95.2) |
| Chronic Cardiac Disease | No | 22615 (66.7) | 4261 (64.8) | 43530 (71.4) | 6571 (69.2) | 15241 (75.5) | 2924 (72.8) | 4004 (68.1) | 595 (68.0) |
|  | Yes | 11298 (33.3) | 2315 (35.2) | 17470 (28.6) | 2918 (30.8) | 4955 (24.5) | 1094 (27.2) | 1874 (31.9) | 280 (32.0) |
| Hypertension | No | 17156 (50.4) | 3495 (53.1) | 32532 (52.9) | 5060 (53.2) | 11717 (57.5) | 2176 (54.1) | 3290 (55.7) | 500 (57.3) |
|  | Yes | 16889 (49.6) | 3083 (46.9) | 28933 (47.1) | 4456 (46.8) | 8674 (42.5) | 1846 (45.9) | 2621 (44.3) | 373 (42.7) |
| Chronic Pulmonary Disease | No | 28521 (84.6) | 4602 (69.9) | 52034 (85.3) | 6752 (70.8) | 17117 (84.5) | 2920 (72.3) | 4736 (80.2) | 581 (65.6) |
|  | Yes | 5189 (15.4) | 1983 (30.1) | 8999 (14.7) | 2784 (29.2) | 3138 (15.5) | 1118 (27.7) | 1169 (19.8) | 305 (34.4) |
| Chronic Renal Disease | No | 27710 (82.3) | 5149 (78.8) | 51719 (85.0) | 7677 (81.1) | 17395 (86.2) | 3132 (78.0) | 4873 (82.7) | 670 (76.4) |
|  | Yes | 5961 (17.7) | 1386 (21.2) | 9136 (15.0) | 1791 (18.9) | 2784 (13.8) | 882 (22.0) | 1020 (17.3) | 207 (23.6) |
| Asthma | No | 29546 (87.7) | 5269 (80.7) | 52021 (85.4) | 7369 (77.5) | 16569 (81.7) | 3138 (77.6) | 4952 (84.0) | 700 (79.5) |
|  | Yes | 4150 (12.3) | 1257 (19.3) | 8922 (14.6) | 2140 (22.5) | 3705 (18.3) | 905 (22.4) | 943 (16.0) | 181 (20.5) |
| Liver Disease | No | 32281 (96.7) | 6204 (95.8) | 58724 (96.9) | 8987 (95.5) | 19463 (96.8) | 3833 (95.8) | 5612 (96.6) | 825 (95.3) |
|  | Yes | 1112 (3.3) | 274 (4.2) | 1856 (3.1) | 422 (4.5) | 635 (3.2) | 168 (4.2) | 197 (3.4) | 41 (4.7) |
| Chronic Neurological Disorder | No | 28925 (86.2) | 5729 (88.1) | 54021 (89.0) | 8478 (89.9) | 18345 (90.9) | 3691 (92.0) | 5049 (86.4) | 779 (89.1) |
|  | Yes | 4640 (13.8) | 776 (11.9) | 6678 (11.0) | 953 (10.1) | 1844 (9.1) | 321 (8.0) | 795 (13.6) | 95 (10.9) |
| Malignant Neoplasm | No | 31019 (92.8) | 4857 (74.6) | 56420 (93.0) | 7206 (76.2) | 18895 (93.8) | 3048 (76.2) | 5269 (90.5) | 628 (71.4) |
|  | Yes | 2423 (7.2) | 1656 (25.4) | 4256 (7.0) | 2252 (23.8) | 1250 (6.2) | 953 (23.8) | 555 (9.5) | 252 (28.6) |
| Chronic Haematologic Disease | No | 32397 (96.9) | 5796 (89.4) | 58907 (97.1) | 8596 (91.2) | 19535 (96.9) | 3600 (89.6) | 5592 (95.9) | 766 (87.8) |
|  | Yes | 1039 (3.1) | 690 (10.6) | 1761 (2.9) | 829 (8.8) | 623 (3.1) | 416 (10.4) | 242 (4.1) | 106 (12.2) |
| Obesity | No | 26042 (87.3) | 5016 (87.5) | 44460 (82.9) | 6693 (81.8) | 14327 (79.0) | 2886 (82.0) | 4728 (87.3) | 705 (87.0) |
|  | Yes | 3793 (12.7) | 717 (12.5) | 9167 (17.1) | 1489 (18.2) | 3815 (21.0) | 634 (18.0) | 687 (12.7) | 105 (13.0) |
| Diabetes | No | 23631 (69.0) | 4669 (70.3) | 43534 (70.7) | 6874 (71.7) | 14424 (70.6) | 2915 (71.8) | 4255 (71.9) | 677 (76.2) |
|  | Yes | 10602 (31.0) | 1970 (29.7) | 18061 (29.3) | 2716 (28.3) | 5995 (29.4) | 1145 (28.2) | 1664 (28.1) | 211 (23.8) |
| Rheumatologic Disorder | No | 29815 (89.5) | 5105 (78.8) | 53567 (88.7) | 7351 (77.9) | 18101 (90.1) | 3116 (77.5) | 5279 (90.3) | 701 (79.8) |
|  | Yes | 3487 (10.5) | 1374 (21.2) | 6858 (11.3) | 2084 (22.1) | 1992 (9.9) | 905 (22.5) | 564 (9.7) | 177 (20.2) |
| Dementia | No | 26981 (80.4) | 5772 (88.7) | 53735 (89.0) | 8762 (93.3) | 18717 (93.1) | 3834 (95.9) | 5130 (87.9) | 837 (94.9) |
|  | Yes | 6588 (19.6) | 735 (11.3) | 6651 (11.0) | 629 (6.7) | 1380 (6.9) | 165 (4.1) | 709 (12.1) | 45 (5.1) |
| Malnutrition | No | 30161 (97.0) | 5841 (97.0) | 55152 (97.9) | 8391 (97.1) | 18759 (98.8) | 3696 (98.7) | 5459 (98.3) | 821 (98.0) |
|  | Yes | 929 (3.0) | 181 (3.0) | 1164 (2.1) | 249 (2.9) | 227 (1.2) | 47 (1.3) | 97 (1.7) | 17 (2.0) |
| Smoking | No | 9736 (52.1) | 1741 (44.1) | 18467 (52.5) | 2578 (45.1) | 5785 (47.8) | 1082 (44.5) | 1390 (43.2) | 186 (35.6) |
|  | Yes | 8969 (47.9) | 2206 (55.9) | 16682 (47.5) | 3132 (54.9) | 6319 (52.2) | 1352 (55.5) | 1828 (56.8) | 336 (64.4) |
| Vaccination Dose on Admission | Unvaccinated | 34396 (100.0) | 6688 (100.0) | 59598 (94.9) | 9143 (94.4) | 13488 (49.7) | 1058 (24.5) | 6882 (93.1) | 848 (92.0) |
|  | First Dose | 0 (0.0) | 0 (0.0) | 3116 (5.0) | 532 (5.5) | 1948 (7.2) | 253 (5.8) | 48 (0.6) | 4 (0.4) |
|  | Second Dose | 0 (0.0) | 0 (0.0) | 55 (0.1) | 11 (0.1) | 11251 (41.5) | 2762 (63.8) | 295 (4.0) | 31 (3.4) |
|  | Third Dose | 0 (0.0) | 0 (0.0) | 0 (0.0) | 0 (0.0) | 428 (1.6) | 251 (5.8) | 164 (2.2) | 39 (4.2) |
|  | Fourth Dose | 0 (0.0) | 0 (0.0) | 0 (0.0) | 0 (0.0) | 2 (0.0) | 2 (0.0) | 0 (0.0) | 0 (0.0) |

**Table S2.** Disease severity by immune status and pandemic wave. The 4C mortality score is calculated on admission to hosptial from age, sex, number of comorbidities, respiratory rate, oxygen saturation, Glasgow coma scale, urea and CRP. The score ranges from 0 (low risk) to 21 (very high risk). Score 0-3 = low risk (1.2% mortality), 4-8 = intermediate risk (9.9% mortality), 9-14 = high risk (31.4% mortality) and 15-21 = very high risk (61.5% mortality).

| label | levels | Immunocompetent - Wave 1 | Immunocompromised - Wave 1 | Immunocompetent - Wave 2 | Immunocompromised - Wave 2 | Immunocompetent - Wave 3 | Immunocompromised - Wave 3 | Immunocompetent - Wave 4 | Immunocompromised - Wave 4 |
| --- | --- | --- | --- | --- | --- | --- | --- | --- | --- |
| ISARIC4C Mortality Score | Median (IQR) | 11.0 (8.0 to 13.0) | 11.0 (9.0 to 13.0) | 10.0 (7.0 to 13.0) | 11.0 (8.0 to 13.0) | 8.0 (4.0 to 11.0) | 10.0 (7.0 to 12.0) | 9.0 (4.0 to 12.0) | 10.0 (7.0 to 12.0) |
| Respiratory Rate (breaths per min) | <20 | 12159 (35.1) | 2259 (33.6) | 20832 (33.1) | 3060 (31.6) | 8563 (32.5) | 1291 (30.3) | 2453 (42.6) | 283 (33.0) |
|  | 20-30 | 16963 (48.9) | 3382 (50.4) | 33071 (52.5) | 5173 (53.5) | 14210 (54.0) | 2406 (56.5) | 2726 (47.4) | 463 (54.0) |
|  | >=30 | 5558 (16.0) | 1074 (16.0) | 9102 (14.4) | 1439 (14.9) | 3545 (13.5) | 564 (13.2) | 578 (10.0) | 112 (13.1) |
| Number of comorbidities | 0 | 4378 (12.3) | 0 (0.0) | 10230 (15.8) | 0 (0.0) | 8238 (30.4) | 0 (0.0) | 1762 (23.8) | 0 (0.0) |
|  | 1 | 6868 (19.3) | 285 (4.2) | 13649 (21.1) | 489 (5.0) | 5809 (21.4) | 335 (7.7) | 1458 (19.7) | 44 (4.8) |
|  | 2+ | 24279 (68.3) | 6573 (95.8) | 40688 (63.0) | 9359 (95.0) | 13070 (48.2) | 3991 (92.3) | 4169 (56.4) | 878 (95.2) |
| Oxygen Saturation (%) | >=92 | 27589 (79.2) | 5254 (77.7) | 49645 (78.6) | 7421 (76.6) | 21201 (80.0) | 3323 (77.9) | 4912 (84.9) | 671 (77.7) |
|  | <92 | 7244 (20.8) | 1507 (22.3) | 13495 (21.4) | 2262 (23.4) | 5288 (20.0) | 942 (22.1) | 875 (15.1) | 193 (22.3) |
| Glasgow Coma Score | 15 | 27882 (84.5) | 5664 (88.2) | 55467 (90.9) | 8585 (91.8) | 24134 (93.9) | 3872 (94.0) | 5403 (91.0) | 771 (92.2) |
|  | <15 | 5128 (15.5) | 756 (11.8) | 5534 (9.1) | 767 (8.2) | 1562 (6.1) | 249 (6.0) | 533 (9.0) | 65 (7.8) |
| Blood Urea (mmol/L) | <7 | 13721 (48.4) | 2633 (47.2) | 28504 (54.9) | 4109 (50.9) | 14805 (66.2) | 1877 (51.3) | 3024 (61.3) | 378 (51.5) |
|  | 7-14 | 9659 (34.0) | 2027 (36.3) | 17073 (32.9) | 2927 (36.3) | 5636 (25.2) | 1291 (35.3) | 1355 (27.5) | 258 (35.1) |
|  | >14 | 4998 (17.6) | 919 (16.5) | 6320 (12.2) | 1032 (12.8) | 1927 (8.6) | 490 (13.4) | 557 (11.3) | 98 (13.4) |
| C-Reactive Protein (mg/L) | <50 | 10342 (35.9) | 1932 (33.9) | 18415 (35.5) | 2658 (32.8) | 8349 (36.8) | 1261 (33.9) | 2552 (52.8) | 307 (42.1) |
|  | 50-99 | 6632 (23.0) | 1400 (24.6) | 13312 (25.7) | 2160 (26.7) | 5556 (24.5) | 1005 (27.0) | 892 (18.5) | 167 (22.9) |
|  | >=100 | 11824 (41.1) | 2363 (41.5) | 20110 (38.8) | 3277 (40.5) | 8757 (38.6) | 1455 (39.1) | 1385 (28.7) | 256 (35.1) |

**Table S3.**
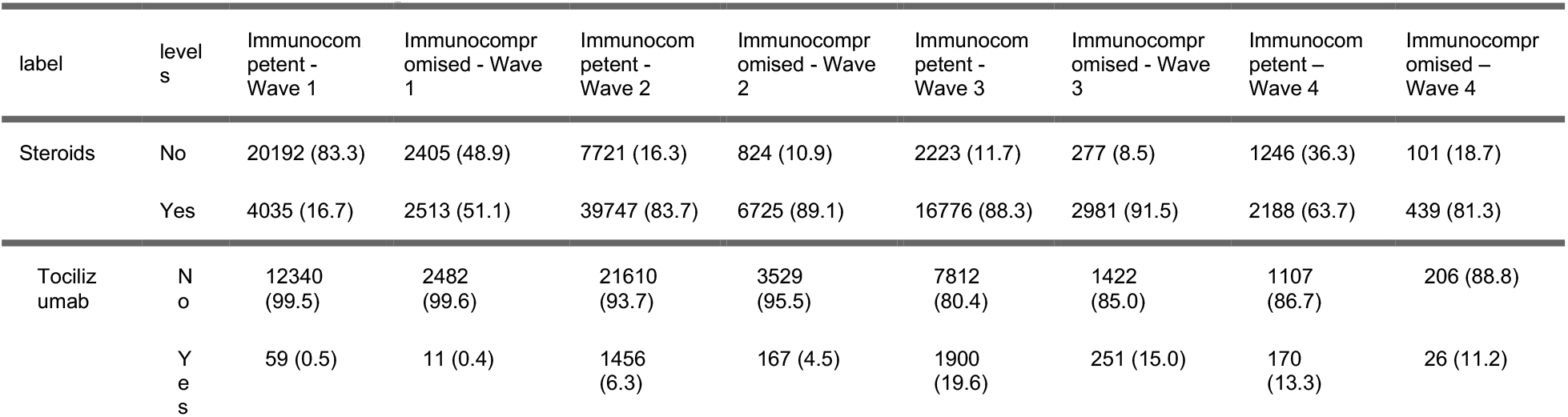
Number (%) of patients receiving steroids and tocilizumab by immune status and pandemic wave.

| label | levels | Immunocompetent - Wave 1 | Immunocompromised - Wave 1 | Immunocompetent - Wave 2 | Immunocompromised - Wave 2 | Immunocompetent - Wave 3 | Immunocompromised - Wave 3 | Immunocompetent - Wave 4 | Immunocompromised - Wave 4 |
| --- | --- | --- | --- | --- | --- | --- | --- | --- | --- |
| Steroids | No | 20192 (83.3) | 2405 (48.9) | 7721 (16.3) | 824 (10.9) | 2223 (11.7) | 277 (8.5) | 1246 (36.3) | 101 (18.7) |
|  | Yes | 4035 (16.7) | 2513 (51.1) | 39747 (83.7) | 6725 (89.1) | 16776 (88.3) | 2981 (91.5) | 2188 (63.7) | 439 (81.3) |
| Tocilizumab | No | 12340 (99.5) | 2482 (99.6) | 21610 (93.7) | 3529 (95.5) | 7812 (80.4) | 1422 (85.0) | 1107 (86.7) | 206 (88.8) |
|  | Yes | 59 (0.5) | 11 (0.4) | 1456 (6.3) | 167 (4.5) | 1900 (19.6) | 251 (15.0) | 170 (13.3) | 26 (11.2) |

**Table S4.** Outcomes by reason for immunocompromise.

| label | levels | Immunocompetent | Pre-Existing Immunological Disorder | Previous Organ Transplant | Cancer | HIV/AIDS | Pre-Admission Immunosuppressants | Pre-Admission Steroids |
| --- | --- | --- | --- | --- | --- | --- | --- | --- |
| Oxygen | No | 38776 (29.2) | 151 (29.2) | 471 (30.6) | 1555 (30.7) | 142 (29.0) | 2858 (22.6) | 350 (22.7) |
|  | Yes | 94128 (70.8) | 367 (70.8) | 1070 (69.4) | 3516 (69.3) | 348 (71.0) | 9769 (77.4) | 1195 (77.3) |
| Critical Care Admission | No | 112738 (84.5) | 421 (81.1) | 1209 (78.2) | 4437 (87.3) | 367 (74.1) | 10637 (84.1) | 1319 (85.2) |
|  | Yes | 20710 (15.5) | 98 (18.9) | 337 (21.8) | 648 (12.7) | 128 (25.9) | 2005 (15.9) | 229 (14.8) |
| Non-Invasive Ventilation | No | 107620 (81.5) | 416 (80.5) | 1170 (76.2) | 4127 (81.8) | 369 (75.8) | 9758 (77.7) | 1212 (79.5) |
|  | Yes | 24445 (18.5) | 101 (19.5) | 365 (23.8) | 918 (18.2) | 118 (24.2) | 2796 (22.3) | 313 (20.5) |
| Invasive Ventilation | No | 123531 (93.4) | 471 (90.9) | 1365 (88.9) | 4820 (95.3) | 428 (87.9) | 11672 (92.9) | 1415 (92.4) |
|  | Yes | 8665 (6.6) | 47 (9.1) | 171 (11.1) | 236 (4.7) | 59 (12.1) | 898 (7.1) | 116 (7.6) |
| Death | No | 102207 (78.1) | 391 (75.9) | 1100 (72.8) | 3168 (63.5) | 396 (81.6) | 8849 (71.4) | 1005 (66.0) |
|  | Yes | 28608 (21.9) | 124 (24.1) | 411 (27.2) | 1818 (36.5) | 89 (18.4) | 3540 (28.6) | 517 (34.0) |

**Figure S1:**
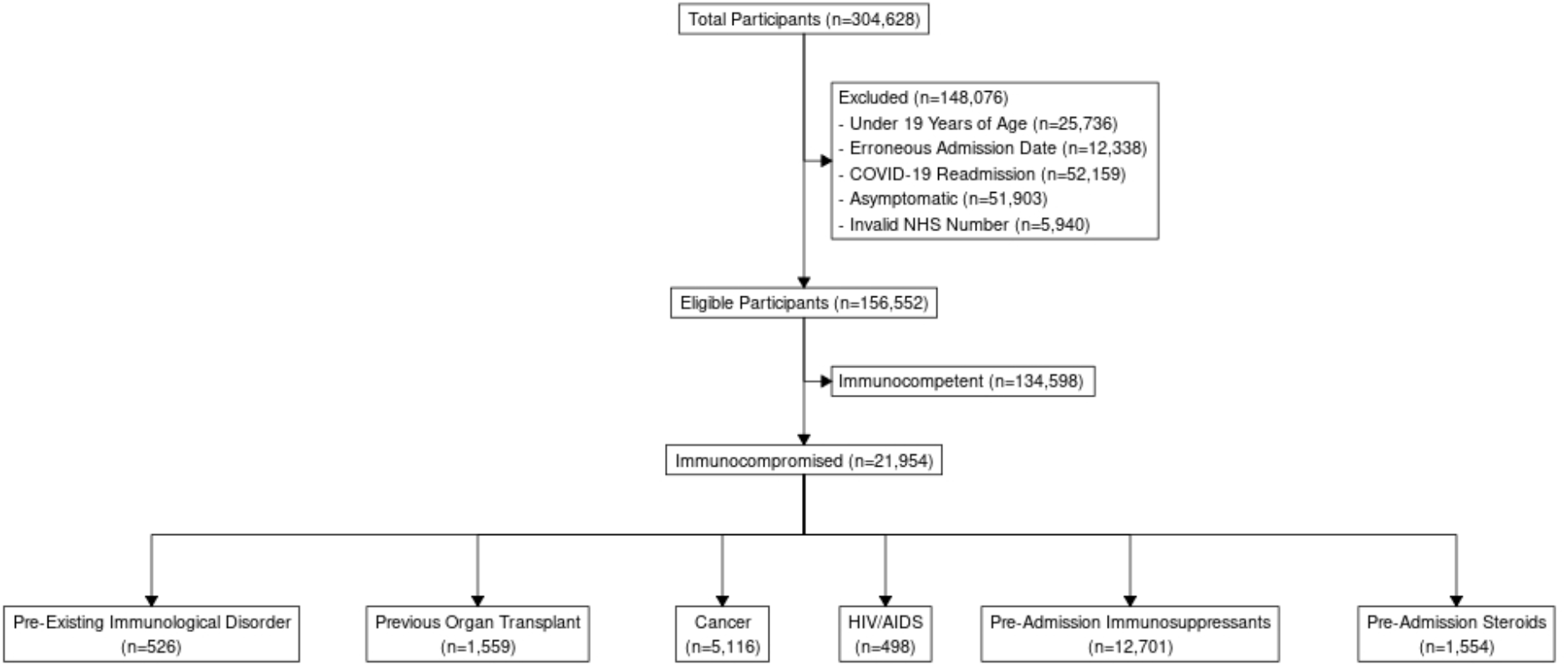
Consort diagram.

**Figure S2.**
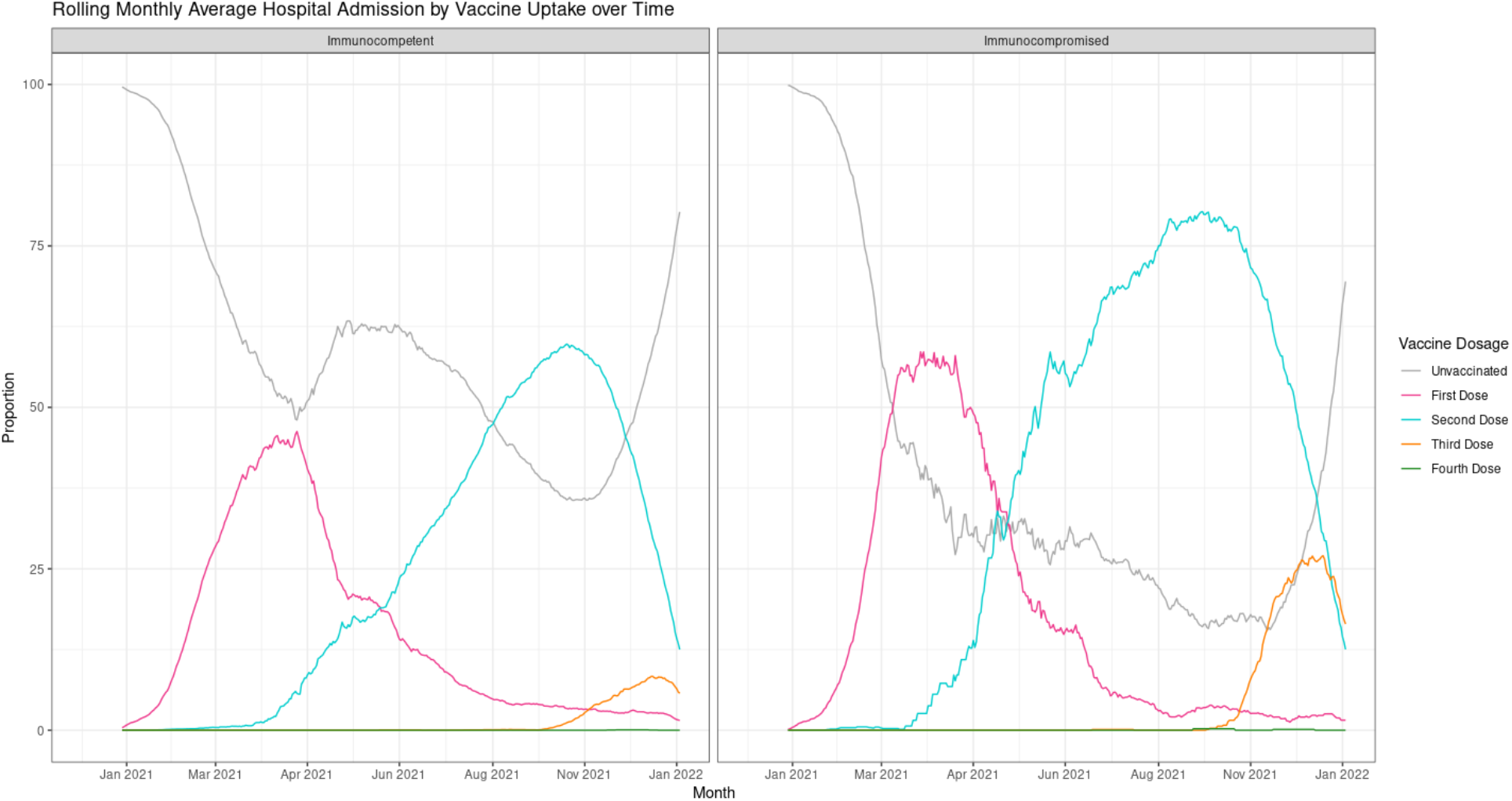
Admissions stratified by the number of vaccine doses received.

**Figure S3.**
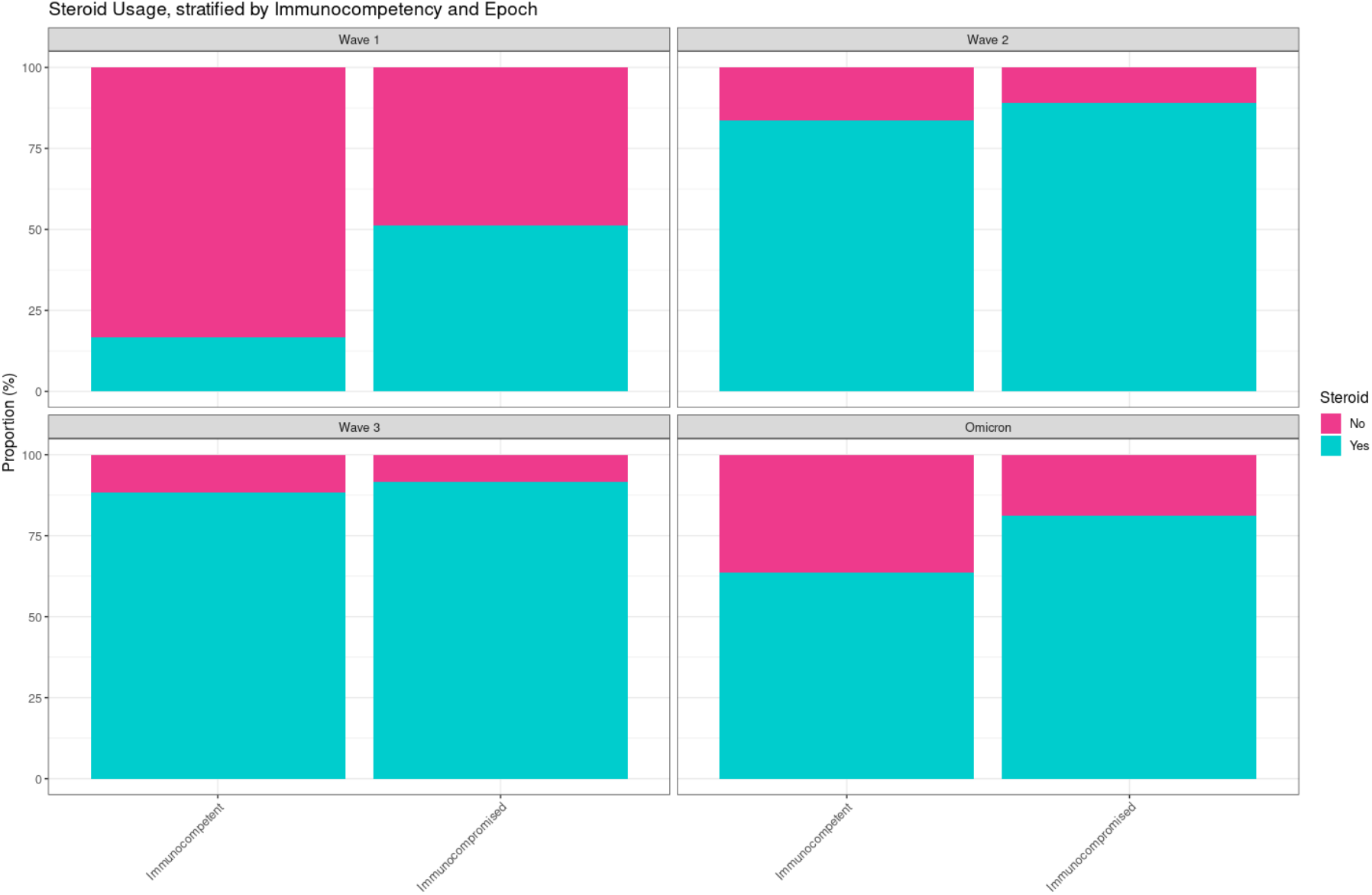
Steroid usage by immune status in the first 4 pandemic waves in the UK.

**Figure S4.**
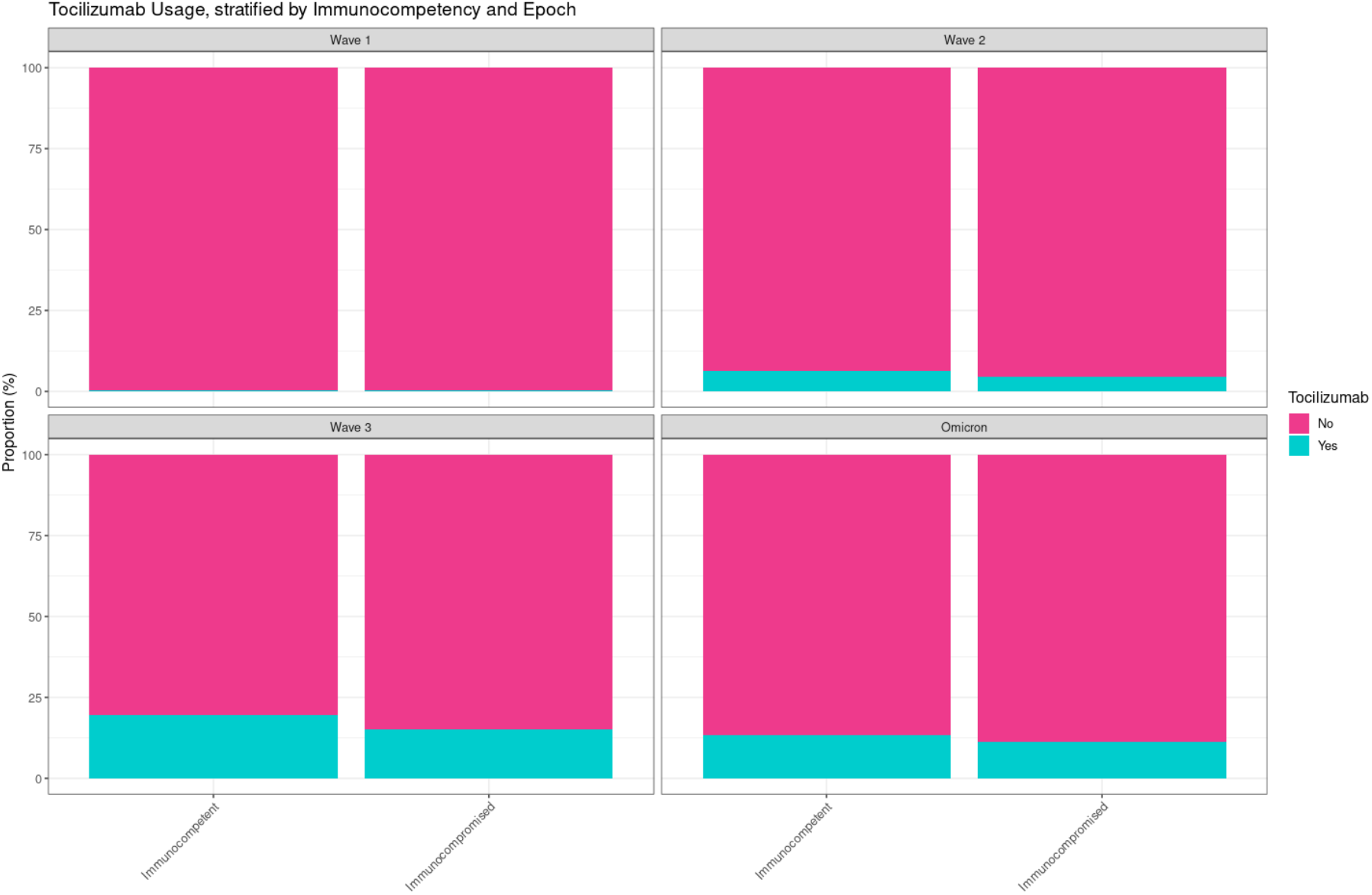
Tocilizumab usage by immune status in the first 4 pandemic waves in the UK.

**Figure S5.**
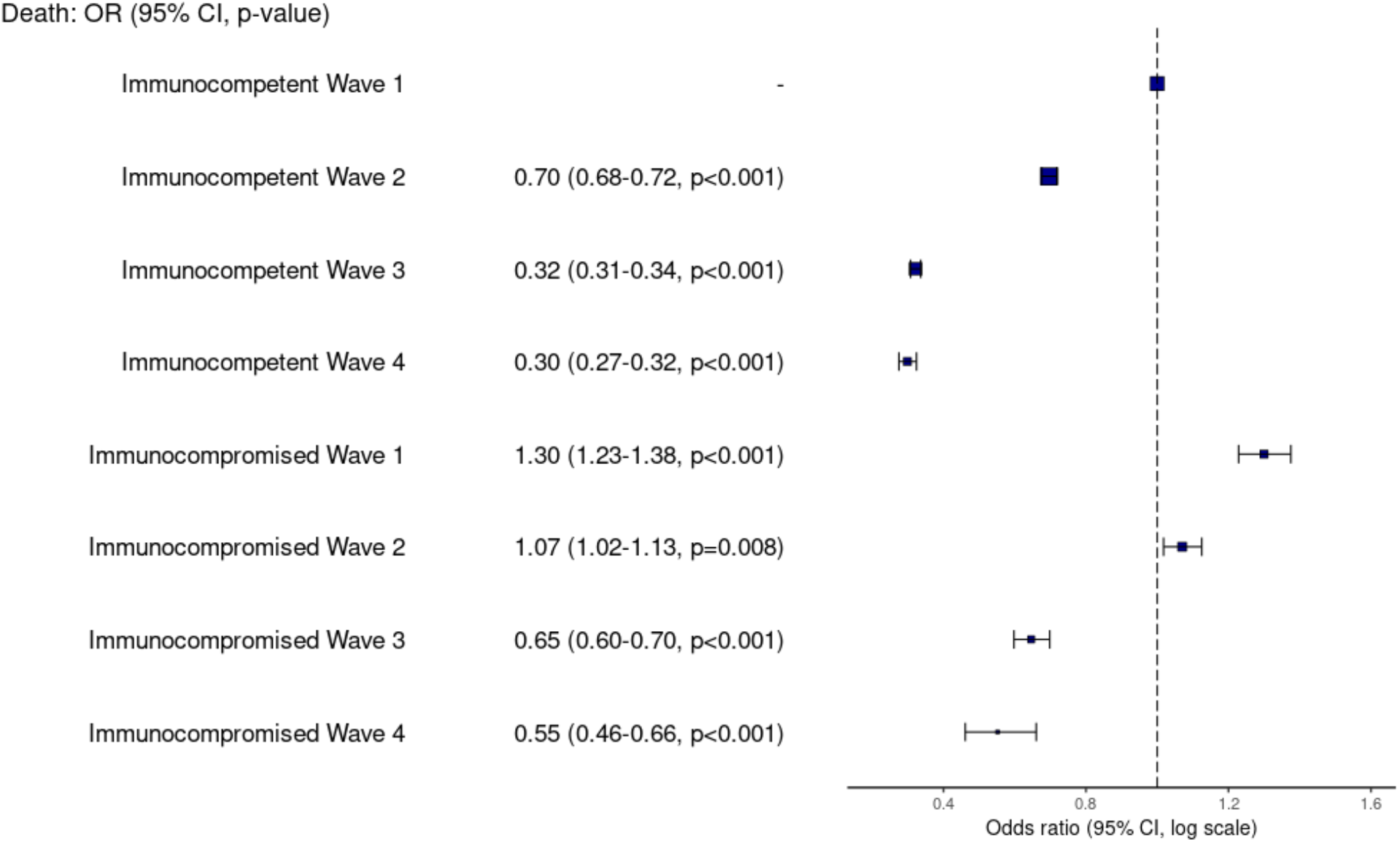
Outcome of hospitalised immunocompromised patients, compared with immunocompetent patients. Odds ratios (OR) for in-hospital death from univariable logistic regression in each of the first four pandemic waves in the UK.

**Figure S6.**
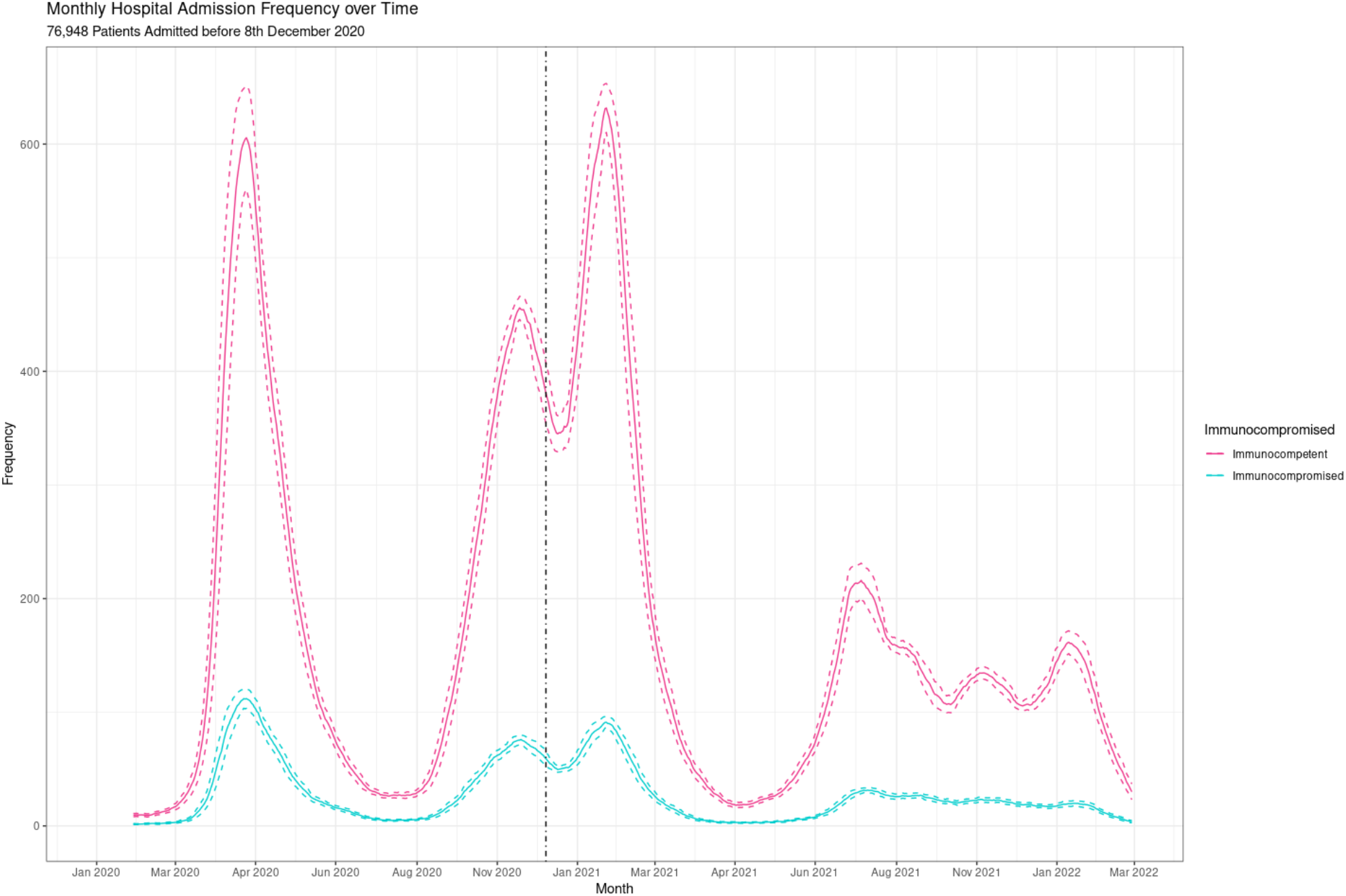
Number of admissions of immunocompetent and immunocompromised patients over time.

